# SARS-CoV-2 Viral Load in Saliva Rises Gradually and to Moderate Levels in Some Humans

**DOI:** 10.1101/2020.12.09.20239467

**Authors:** Alexander Winnett, Matthew M. Cooper, Natasha Shelby, Anna E. Romano, Jessica A. Reyes, Jenny Ji, Michael K. Porter, Emily S. Savela, Jacob T. Barlow, Reid Akana, Colten Tognazzini, Matthew Feaster, Ying-Ying Goh, Rustem F. Ismagilov

## Abstract

Transmission of SARS-CoV-2 in community settings often occurs before symptom onset, therefore testing strategies that can reliably detect people in the early phase of infection are urgently needed. Early detection of SARS-CoV-2 infection is especially critical to protect vulnerable populations who require frequent interactions with caretakers. Rapid COVID-19 tests have been proposed as an attractive strategy for surveillance, however a limitation of most rapid tests is their low sensitivity. Low-sensitivity tests are comparable to high sensitivity tests in detecting early infections when two assumptions are met: (1) viral load rises quickly (within hours) after infection and (2) viral load reaches and sustains high levels (>10^5^– 10^6^ RNA copies/mL). However, there are no human data testing these assumptions. In this study, we document a case of presymptomatic household transmission from a healthy young adult to a sibling and a parent. Participants prospectively provided twice-daily saliva samples. Samples were analyzed by RT-qPCR and RT-ddPCR and we measured the complete viral load profiles throughout the course of infection of the sibling and parent. This study provides evidence that in at least some human cases of SARS-CoV-2, viral load rises slowly (over days, not hours) and not to such high levels to be detectable reliably by any low-sensitivity test. Additional viral load profiles from different samples types across a broad demographic must be obtained to describe the early phase of infection and determine which testing strategies will be most effective for identifying SARS-CoV-2 infection before transmission can occur.

**One sentence summary:** In some human infections, SARS-CoV-2 viral load rises slowly (over days) and remains near the limit of detection of rapid, low-sensitivity tests.

## Introduction

As of early December 2020, nearly one year after the first COVID-19 outbreak in Wuhan, China, there have been more than 65 million cases and 1.5 million deaths globally.^1^ Transmission of SARS-CoV-2 in community settings often occurs before symptom onset,^2,3^ putting at great risk people who require frequent interactions with caregivers, such as residents of nursing homes. Better strategies for using the available COVID-19 diagnostic tests are critically needed to decrease overall transmission, thereby reducing transmission to these vulnerable populations.^4^

Transmission from asymptomatic or presymptomatic individuals is considered the Achilles’ Heel of COVID-19 infection control.^3^ In a recent epidemiologic investigation of 183 confirmed COVID-19 cases in Wanzhou, China, about 76% of transmissions occurred from individuals without symptoms (either asymptomatic or presymptomatic).^5^ Numerous transmission events originating from individuals without symptoms have been documented in a variety of locations, such as dinner parties,^6^ skilled nursing facilities,^2,7^ correctional facilities,^8^ sporting events,^9,10^ religious ceremonies,^6^ and spring break trips.^11^

The importance of effective testing strategies to quell transmission from individuals without symptoms is underscored by an outbreak at an overnight camp in Wisconsin^12^ where a 9^th^ grade student developed symptoms the day after arrival, prompting quarantine of 11 close contacts. All 11 contacts were asymptomatic and released from quarantine after receiving negative rapid antigen test results. However, 6 of the 11 went on to develop symptoms, and an outbreak ensued, with more than 100 additional individuals (a total of 3/4 of camp attendees) infected. In contrast, a more successful containment of an outbreak was documented in a skilled nursing facility in Los Angeles, where serial surveillance PCR testing was initiated immediately after three residents became symptomatic and tested positive.^13^ PCR results prompted isolation of 14 infected individuals without symptoms, which limited the outbreak to a total of only 19 out of 99 residents over the course of two weeks. These cases demonstrate the value of testing strategies that can detect and isolate infected individuals in the early phase in the infection, reducing the potential for transmission to others during the elicitation window (the period when a person is infectious but not isolating).^14^

More than 200 *in vitro* diagnostics have received Emergency Use Authorization (EUA) from the U.S. Food and Drug Administration for identification of acute SARS-CoV-2 infection.^15^ These tests have a wide range of sensitivities. The most sensitive tests, with limits of detection (LOD) of 10^2^-10^3^ RNA copies/mL, include the RT-qPCR assays. These tests typically involve more intensive sample-preparation methods to extract and purify RNA and most are run in centralized laboratories (with a few exceptions of point-of-care tests that integrate rigorous sample preparation and RNA detection^15,16^). At the other extreme are the low-sensitivity tests (LODs of ∼10^5^–10^7^ RNA copies/mL), such as antigen tests or molecular tests that do not perform rigorous sample preparation. These tests offer tangible advantages, such as being fast (rapid antigen tests yield results in minutes), less expensive to manufacture, and can be deployable outside of laboratories.

Rapid, low-sensitivity tests are clearly a valuable part of the overall infection control strategy; however, the use of such tests as a strategy for diagnosing infected persons at the early phase of infection is controversial. The U.S. Food & Drug Administration (FDA) has authorized such tests for use in symptomatic populations. Data in several reports suggest that such tests may miss presymptomatic and asymptomatic individuals early in the infection.^12,17^ However, logical arguments have also been made,^18,19^ in favor of widely deploying surveillance tests with “analytic sensitivities vastly inferior to those of benchmark tests.”^18^

Low-sensitivity tests will be equally effective to high-sensitivity tests at minimizing transmission if the following two assumptions about the early phase of SARS-CoV-2 infection hold true: (i) viral load increases rapidly, by orders of magnitude within hours, and (ii) viral load reaches and sustains high levels during the infectious window, such that a rapid low-sensitivity test would have a similar ability to detect early-phase infections compared with high-sensitivity tests. These two assumptions have not been tested in humans. Viral load at the early phases of SARS-CoV-2 infection remains a knowledge gap necessary to inform the use of testing resources to effectively minimize transmission. To fill this knowledge gap, and inform selection of diagnostic tests appropriate for identifying infections in the earliest phases, requires studies that monitor SARS-CoV-2 viral load with high temporal resolution (beginning at the incidence of infection) and in a large, diverse cohort of individuals.

We are conducting a case-ascertained observational study in which community members recently diagnosed with COVID-19 and their SARS-CoV-2-presumed-negative household contacts prospectively provide twice-daily saliva samples. We are quantifying absolute SARS-CoV-2 RNA viral load from these saliva samples using RT-qPCR and RT digital droplet PCR (RT-ddPCR) assays. This article documents preliminary results from the study, with the complete SARS-Cov-2 viral load profiles from two cases of observed household transmission.

## Methods

### Participant Population

This study was reviewed and approved by the Institutional Review Board of the California Institute of Technology, protocol #20-1026. All participants provided written informed consent prior to participation. Individuals ages 6 and older were eligible for participation if they lived within the jurisdiction of a partnering public health department and were recently (within 7 days) diagnosed with COVID-19 by a CLIA laboratory test or were currently living in a shared residence with at least one person who was recently (within 7 days) diagnosed with COVID-19 by a CLIA laboratory test. The exclusion criteria for the study included physical or cognitive impairments that would affect the ability to provide informed consent, or to safely self-collect and return samples. In addition, participants must not have been hospitalized, and they must be fluent in either Spanish or English. Individuals without laboratory confirmed COVID-19 but with symptoms of respiratory illness in the 14 days preceding screening for enrollment were not eligible. Study data were collected and managed using REDCap (Research Electronic Data Capture) hosted at the California Institute of Technology.

### Symptom Monitoring

Participants in the study completed a questionnaire upon enrollment to provide information on demographics, health factors, COVID-19 diagnosis history, COVID-19-like symptoms since February 2020, household infection-control practices and perceptions of COVID-19 risk. Additionally, participants recorded any COVID-19-like symptoms (as defined by the U.S. Centers for Disease Control^20^) that they were experiencing on a symptom-tracking card at least once per day. Participants also filled out an additional questionnaire at the conclusion of the study to document behaviors and interactions with household members during their enrollment.

### Collection of Respiratory Specimens

Participants self-collected saliva samples using the Spectrum SDNA-1000 Saliva Collection Kit (Spectrum Solutions LLC, Draper, UT, USA) at home twice per day (after waking up and before going to bed), following the manufacturer’s guidelines. Participants were instructed not to eat, drink, smoke, brush their teeth, use mouthwash, or chew gum for at least 30 min prior to donating. These tubes were labelled and packaged by the participants and transported at room temperature by a medical courier to the California Institute of Technology daily for analysis.

### Nucleic Acid Extraction

An aliquot of 400 µL from each saliva sample in Spectrum buffer was manually extracted using the MagMAX Viral/Pathogen Nucleic Acid Isolation Extraction Kit (Cat. A42352, Thermo Fisher Scientific) and eluted in 100 µL. Positive extraction controls and negative extraction controls were included in every extraction batch: Positive extraction controls were prepared by combining 200 µL commercial pooled human saliva (Cat. 991-05-P, Lee Bioscience) with 200 µL SARS-CoV-2 heat inactivated particles (Cat. NR-52286, BEI Resources) at a concentration equivalent to 7500 genomic equivalent units/mL, mixed with the buffer from the Spectrum SDNA-1000 Saliva Collection Device. Negative extraction controls were prepared by combining 200 µL commercial pooled human saliva and 200 µL Spectrum buffer. Spectrum buffer contained components that inactivate the virus and stabilize the RNA, facilitating the study.

### Quantification of Viral Load by RT-qPCR

An aliquot of 5 µL of eluent was input into duplicate 20 µL RT-qPCR reactions (Cat. A15299, TaqPath 1-Step RT-qPCR Master Mix, CG) with multiplex primers and probes from Integrated DNA Technologies (Coralville, IA, USA) targeting SARS-CoV-2 N1 (Cat. 10006821, 10006822, 10006823) and N2 (Cat. 10006824, 10006825, 10007050), and human RNase P (Cat. 10006827, 10006828, 10007061). Positive and negative reaction controls were included on every plate: Templates for positive control reactions contained 4 copies per µL of SARS-CoV-2 genomic RNA from nCoV 2019-nCoV/USA-WA1/2020 (Cat.

NR-52281, BEI) in 5 µL of nuclease-free water (Cat. AM9932, ThermoFisher Scientific), and negative reaction controls contained only nuclease-free water. Reactions were run on a CFX96 Real-time PCR System (Bio-Rad Laboratories) according to the amplification protocol defined in the CDC 2019-Novel Coronavirus (2019-nCoV) Real-Time RT-PCR Diagnostic Panel, with Ct determination by auto-thresholding each target channel. Ct values were converted to viral load relative to the resulting value of known input of SARS-CoV-2 heat-inactivated particles in the positive extraction control. The conversion was done across all qPCR samples, using the average Ct values of the positive control for the two SARS-CoV-2 gene targets (N1 and N2; each 32.50 and N=11) after auto-thresholding on cycles 10-45.

### Quantification of Viral Load by RT-ddPCR

An aliquot of 5.5 µL from a dilution of the eluent (samples were diluted to be within the range required for ddPCR) was input into 22 µL reactions of the Bio-Rad SARS-CoV-2 ddPCR Kit (Cat. 12013743, BioRad Laboratories) for multiplex quantification of SARS-CoV-2 N1 and N2 targets, and human RNase P targets. Droplets were generated on a QX200 droplet generator (#1864002, Bio-Rad Laboratories) and measured using a QX200 Droplet Digital PCR System (#1864001, Bio-Rad Laboratories), with analysis using QuantaSoft Analysis Software.

### Conversion of Ct Values to Viral Load

RT-qPCR Ct values from our assay were converted to viral load (copies/mL) using the following equation:

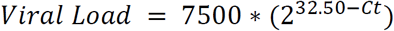

Both the CLIA Laboratory and the laboratory analysis in Kissler et al.^21^ utilized similar assays to ours, and therefore we assumed this same equation could be used to estimate viral load from Ct values from those sources.

### Sequencing

Extracted RNA from samples taken from the early, peak, and late infection stages of each of the three individuals infected with SARS-CoV-2 was sequenced by the Chan Zuckerberg Biohub (San Francisco, CA, USA). All sequences, throughout infection, and across household contacts, were found to be identical.

## Results and Discussion

We report a case of SARS-CoV-2 transmission in a household of four individuals, who we refer to as Parent-1, Parent-2, Sibling-1, and Sibling-2 (Table S1). Sibling-1 (who reported recent close contact with someone infected with SARS-CoV-2) and Sibling-2 returned home together from out-of-state and were CLIA-lab RT-qPCR tested for COVID-19 the next day. The following day, Sibling-2’s specimen resulted negative, and Sibling-1’s specimen resulted positive, prompting Sibling-1 to isolate and all household members to quarantine. Within hours of receiving Sibling-1’s positive-test result, they were enrolled in the study. Parent-2 remained SARS-CoV-2-negative in all samples. Parent-1 and Sibling-2 were SARS-CoV-2-negative upon enrollment and became continuously positive starting ∼36 hours after enrollment. Viral sequencing determined that the SARS-CoV-2 in samples from Sibling-1, Parent-1 and Sibling-2 shared identical sequences to each other, highly supportive of household transmission.

All nucleic acid measurements from saliva samples included human RNase P target measurements (Figure S2) as an indicator of sample quality to confirm that viral load dynamics were not an artifact of sample collection. RNase P Ct values were consistent across samples from each participant: during the early phase of infection (from enrollment to just beyond peak viral load, up to day 8 of enrollment), the 15 samples from Parent-1 had an average RNase P Ct value of 27.28 (±1.12 SD), and the 15 samples from Sibling-2 had an average Ct value of 24.51 (±1.31 SD). Also, a pattern of lower RNase P Ct values (more human material) in morning samples than evening samples is occasionally discernable, although this pattern does not appear to dominate viral load signal.

RT-qPCR and RT-ddPCR measurements of viral load from saliva samples provided by the three infected individuals (Figure 1) offer three insights. (i) Presymptomatic viral loads in some humans can rise over the course of days not hours, which is slower than expected.^18,22^ This slow rise increases the utility of sensitive tests (such as PCR) to enable earlier detection and isolation of infected individuals before their viral load increases to a presumably more infectious level. Sibling-2’s viral load rose slowly—within PCR detection range—for 3 days until viral load reached the limit of detection (LOD) for rapid tests. This slow rise also makes it more dangerous to assume that most persons with low viral load will not become infectious^19^ and therefore do not need to self-isolate: Sibling-2 produced 6 positive samples in the 10^3^-10^5^ copies/mL range presymptomatically before peaking at ∼10^7^ copies/mL. (ii) Peak viral load does not always rise above the LOD of rapid, low-sensitivity tests, as expected.^18,22^ Of 88 positive SARS-CoV-2 samples, only one sample was well above the LOD range of rapid tests. Furthermore, in Sibling-1, the logical source of the two infections, neither the CLIA-lab test nor our testing detected viral loads above 10^7^ copies/mL during the presumed period of household transmission. (iii) The LOD of a test can affect how early in infection we can diagnose an infected person, and how consistently we can detect early-phase infections. Of the 52 positive samples from the first 10 days of the study, 33 were near or above the LOD of the more sensitive rapid test (ID NOW, as determined by the FDA) whereas only 3 were near or above the least sensitive LOD of 9.3*10^6^ copies/mL (Table S2). Importantly, there were several days during the presymptomatic period that Parent-1 and Sibling-2 were detected by RT-qPCR, but may not have been reliably detected by many low-sensitivity tests.

**FIGURE 1.**
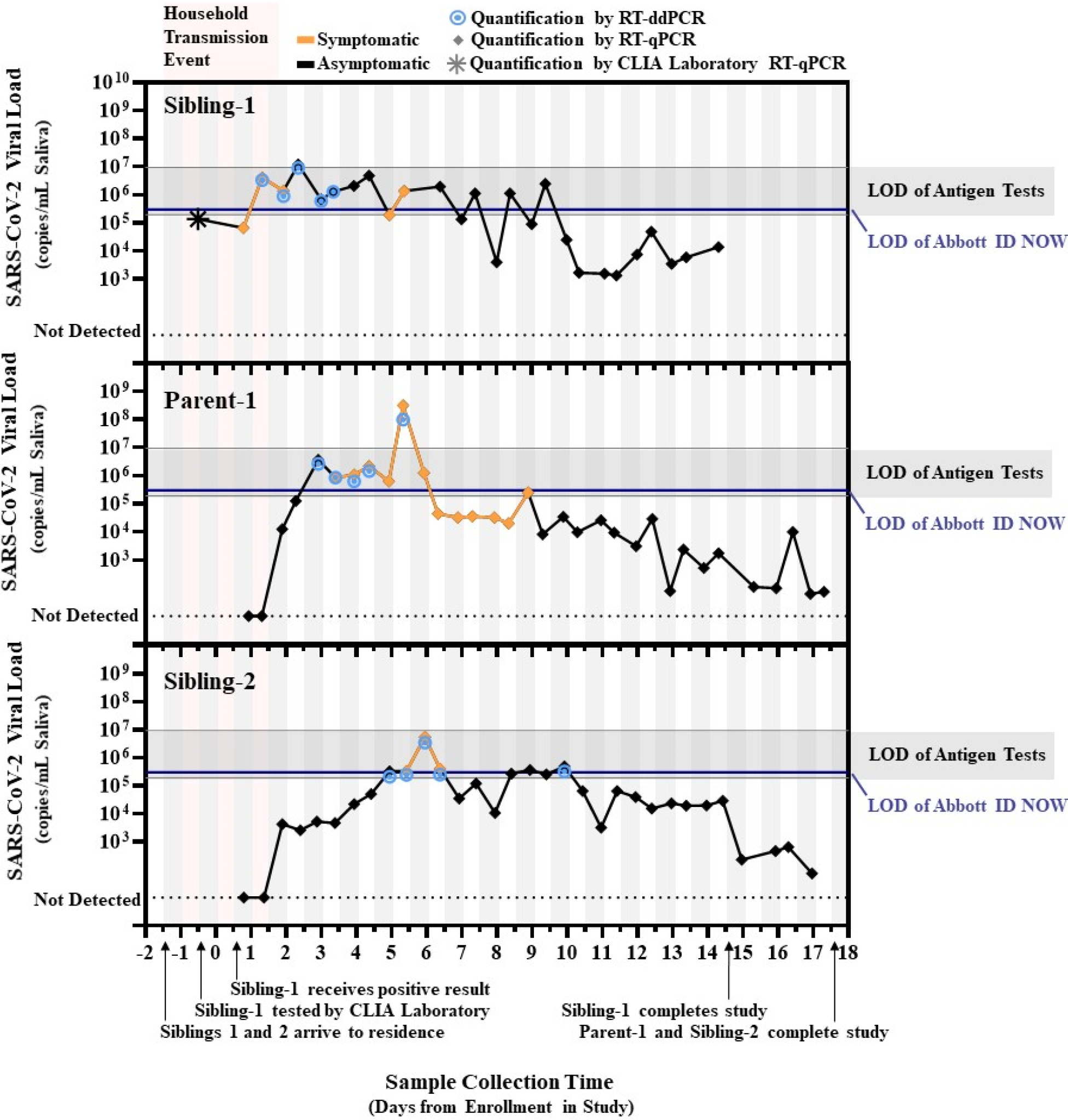
Quantified SARS-CoV-2 saliva viral load after transmission between household contacts relative to detection limits of rapid tests. SARS-CoV-2 viral load over time for “Sibling-1” (the household index case), as well as “Parent-1” and “Sibling-2.” All three viral sequences were identical. Star indicates the viral load estimated from the cycle threshold (Ct) result from the commercial CLIA laboratory test used to diagnose Sibling-1. Diamonds indicate conversion from N1 target cycle threshold values obtained by RT-qPCR to SARS-CoV-2 viral load. Bullseyes indicate viral load obtained by single-molecule RT droplet digital PCR (RT-ddPCR). Black lines represent periods when participants reported no symptoms; orange lines indicate periods when participants reported at least one symptom. Vertical bars indicate the before noon (white) and after noon (grey) periods of each day. Pink shading indicates the presumed period of the household transmission events. Horizontal blue lines depict the limit of detection (LOD) of the Abbott ID NOW (3 × 10^5^ copies/mL) for upper respiratory specimens from U.S. FDA SARS-CoV-2 Reference Panel Comparative testing data. Horizontal grey bars depict the range of LODs estimated for commercial antigen tests for upper respiratory specimens (1.90 × 10^5^ copies/mL to 9.33 × 10^6^ copies/mL; see Table S2).

The insights from this study are also supported by analysis of data by Kissler et al.,^21^ reporting longitudinal (but less frequent) testing of anterior nares and oropharyngeal swabs from individuals associated with the National Basketball Association (Figure S1). In Kissler et al.^21^, viral loads rose slowly (for up to 5 days in some individuals) between the first PCR positive test to the LOD of rapid tests. Few of these samples ever reached viral loads well above the LODs of most rapid antigen tests (Table S2). Low-sensitivity tests have a role in the COVID-19 testing strategies, but our limited data from this study and data from Kissler et al.^21^ demonstrate clearly that for at least some individuals, low-sensitivity tests will likely be unable to reliably diagnose SARS-CoV-2 infection during the early phase of infection. Our limited data are consistent with the use of low-sensitivity tests for point-of-care confirmation of suspected COVID-19 in symptomatic individuals, as authorized by the FDA, but not for universal surveillance testing of asymptomatic individuals, as has been proposed.^18,19,22^

Additional studies are urgently needed to address several limitations of this work. High-frequency viral load measurements from the incidence of infection must be observed in a larger, diverse pool of participants to infer the distribution of viral load profiles in human SARS-CoV-2 infection. Both saliva and nasal swabs have been proposed as sample types for rapid, low-sensitivity tests; however, the LOD of these tests is better defined in nasal swabs. Our study only analyzed saliva; although saliva has been demonstrated to be a more sensitive sample type than nasopharyngeal swab by some studies,^23^ other studies have arrived at the opposite conclusion.^24,25^ The details of saliva collection, sample stabilization, preanalytical handling, sample-preparation protocols, and timing of sampling may play a role in the apparent sensitivity measured in different sample types. No previous study has directly compared saliva with other sample types during the early phase of SARS-CoV-2 infection. Quantitative comparisons of multiple respiratory sample types (including nasal, oropharyngeal, and nasopharyngeal swabs) at the same time points are needed to clarify viral load profiles in different respiratory specimen types. Lastly, to understand the relationship between viral load and infectiousness, direct comparisons of RNA viral load to culturable virus titer across the entire course of infection are needed. Data to address these limitations are needed to inform optimal testing strategies to reduce SARS-CoV-2 transmission.

Understanding the kinetics of viral load from the incidence of infection and throughout the infectious period for a broad demographic will have implications on SARS-CoV-2 testing policies, including policies around travel. For example, several states require recent negative test results for out-of-state visitors prior to arrival. Although some states acknowledge the risk of false negative results by low-sensitivity tests and require PCR confirmation,^19,26^ others do not specify the type of negative test result required for arrival.^27^ If rapid low-sensitivity tests are used, they would risk missing presymptomatic individuals before they rise to their highest (and presumably most infectious) viral load. Such false negative tests have the potential to create a costly false sense of security; individuals who may have been recently exposed and receive a negative result from a low-sensitivity test may be more likely—compared to individuals who did not get a test—to come into contact with other members of the community, including the most vulnerable populations.

All tests have value when used properly and within the right strategy, and we anticipate that these preliminary results will stimulate studies that will provide a better understanding of SARS-CoV-2 viral load in the early stages of infection and lead to the development of effective testing strategies.

## Supporting information

COI forms - all authors

## Data Availability

Raw Ct values and calculated viral loads are available at CaltechDATA, https://data.caltech.edu/records/1702

## COMPETING INTERESTS STATEMENT

The authors report grants from the Bill & Melinda Gates Foundation (to RFI) and the Jacobs Institute for Molecular Engineering for Medicine (to RFI), as well as a Caltech graduate fellowship (to MMC), two National Institutes of Health Biotechnology Leadership Pre-doctoral Training Program (BLP) fellowships (to MKP and JTB) from Caltech’s Donna and Benjamin M. Rosen Bioengineering Center (T32GM112592), and a National Institutes of Health NIGMS Predoctoral Training Grant (GM008042) (to AW) during the course of the study. Dr. Ismagilov is a co-founder, consultant, and a director and has stock ownership and received personal fees from Talis Biomedical Corp., outside the submitted work. In addition, Dr. Ismagilov is an inventor on a series of patents licensed to Bio-Rad Laboratories Inc. in the context of ddPCR, with royalties paid.

## FUNDING

This study is based on research funded in part by the Bill & Melinda Gates Foundation. The findings and conclusions contained within are those of the authors and do not necessarily reflect positions or policies of the Bill & Melinda Gates Foundation. This work was also funded by the Jacobs Institute for Molecular Engineering for Medicine. AW is supported by an NIH NIGMS Predoctoral Training Grant (GM008042); MMC is supported by a Caltech Graduate Student Fellowship; and MP and JTB are each partially supported by National Institutes of Health Biotechnology Leadership Pre-doctoral Training Program (BLP) fellowships from Caltech’s Donna and Benjamin M. Rosen Bioengineering Center (T32GM112592).

## ACKNOWLEDGEMENTS

We thank Lauriane Quenee, Junie Hildebrandt, Grace Fisher-Adams, RuthAnne Bevier, Chantal D’Apuzzo, Ralph Adolphs, Victor Rivera, Steve Chapman, Gary Waters, Leonard Edwards, and Shannon Yamashita for their assistance and advice on study implementation or administration. We thank Jessica Leong, Jessica Slagle, and Angel Navarro for volunteering their time to help with this study. We thank Kissler et al. for making their data publicly available to the community. We thank Maira Phelps, Lienna Chan, Lucy Li and Amy Kistler at the Chan Zuckerberg Biohub for performing SARS-CoV-2 sequencing. We thank Jennifer Fulcher, Debika Bhattacharya and Matthew Bidwell Goetz for their ideas on potential study populations and early study design. We thank Martin Hill, Alma Sanchez, Scott Kim, Debbie Noble, Nina Paddock, Whitney Harrison, and Stu Miller for their support with recruitment. We thank Allison Rhines, Karen Heichman, and Dan Wattendorf for valuable discussion and guidance. Finally, we thank all the Pasadena Public Health Department case investigators and contact tracers for their efforts in study recruitment and their work in the pandemic response.

## SUPPLEMENTARY MATERIALS

Supplemental Tables 1-2

Supplemental Figures 1-2

Supplemental References

Contributions of Non-Corresponding Authors

## Notes

### Author Declarations

This study was reviewed and approved by the Institutional Review Board of the California Institute of Technology, protocol #20-1026.

