## Supplementary material for "SARS-CoV-2 Viral Load in Saliva Rises Gradually and to Moderate Levels in Some Humans": COI forms - all authors

INTERNATIONAL COMMITTEE of MEDICAL JOURNAL EDITORS

### ICMJE Form for Disclosure of Potential Conflicts of Interest

#### Instructions

**The purpose of this form is to provide readers of your manuscript with information about your other interests that could influence how they receive and understand your work. The form is designed to be completed electronically and stored electronically. It contains programming that allows appropriate data display. Each author should submit a separate form and is responsible for the accuracy and completeness of the submitted information. The form is in six parts.**

#### 1. Identifying information.

#### 2. The work under consideration for publication.

This section asks for information about the work that you have submitted for publication. The time frame for this reporting is that of the work itself, from the initial conception and planning to the present. The requested information is about resources that you received, either directly or indirectly (via your institution), to enable you to complete the work. Checking "No" means that you did the work without receiving any financial support from any third party -- that is, the work was supported by funds from the same institution that pays your salary and that institution did not receive third-party funds with which to pay you. If you or your institution received funds from a third party to support the work, such as a government granting agency, charitable foundation or commercial sponsor, check "Yes".

#### 3. Relevant financial activities outside the submitted work.

This section asks about your financial relationships with entities in the bio-medical arena that could be perceived to influence, or that give the appearance of potentially influencing, what you wrote in the submitted work. You should disclose interactions with ANY entity that could be considered broadly relevant to the work. For example, if your article is about testing an epidermal growth factor receptor (EGFR) antagonist in lung cancer, you should report all associations with entities pursuing diagnostic or therapeutic strategies in cancer in general, not just in the area of EGFR or lung cancer.

Report all sources of revenue paid (or promised to be paid) directly to you or your institution on your behalf over the 36 months prior to submission of the work. This should include all monies from sources with relevance to the submitted work, not just monies from the entity that sponsored the research. Please note that your interactions with the work's sponsor that are outside the submitted work should also be listed here. If there is any question, it is usually better to disclose a relationship than not to do so.

For grants you have received for work outside the submitted work, you should disclose support ONLY from entities that could be perceived to be affected financially by the published work, such as drug companies, or foundations supported by entities that could be perceived to have a financial stake in the outcome. Public funding sources, such as government agencies, charitable foundations or academic institutions, need not be disclosed. For example, if a government agency sponsored a study in which you have been involved and drugs were provided by a pharmaceutical company, you need only list the pharmaceutical company.

#### 4. Intellectual Property.

This section asks about patents and copyrights, whether pending, issued, licensed and/or receiving royalties.

#### 5. Relationships not covered above.

Use this section to report other relationships or activities that readers could perceive to have influenced, or that give the appearance of potentially influencing, what you wrote in the submitted work.

##### Definitions.

**Entity:** government agency, foundation, commercial sponsor, academic institution, etc.
**Grant:** A grant from an entity, generally [but not always] paid to your organization
**Personal Fees:** Monies paid to you for services rendered, generally honoraria, royalties, or fees for consulting , lectures, speakers bureaus, expert testimony, employment, or other affiliations
**Non-Financial Support:** Examples include drugs/equipment supplied by the entity, travel paid by the entity, writing assistance, administrative support, etc**.**

**Other:** Anything not covered under the previous three boxes
**Pending:** The patent has been filed but not issued
**Issued:** The patent has been issued by the agency
**Licensed:** The patent has been licensed to an entity, whether earning royalties or not
**Royalties:** Funds are coming in to you or your institution due to your patent

### ICMJE Form for Disclosure of Potential Conflicts of Interest

#### Section 1. Identifying Information

1. Given Name (First Name)
Reid

2. Surname (Last Name)
Akana

3. Date
20-November-2020

4. Are you the corresponding author? ☐ Yes ☑ No

Corresponding Author's Name
Rustem F. Ismagilov

5. Manuscript Title
SARS-CoV-2 Viral Load Rises Gradually and to Moderate Levels in Some Humans

6. Manuscript Identifying Number (if you know it)
n/a

#### Section 2. The Work Under Consideration for Publication

Did you or your institution **at any time** receive payment or services from a third party (government, commercial, private foundation, etc.) for any aspect of the submitted work (including but not limited to grants, data monitoring board, study design, manuscript preparation, statistical analysis, etc.)?

Are there any relevant conflicts of interest? ☑ Yes ☐ No

If yes, please fill out the appropriate information below. If you have more than one entity press the "ADD" button to add a row. Excess rows can be removed by pressing the "X" button.

| Name of Institution/Company | Grant? | Personal Fees? | Non-Financial Support? | Other? | Comments |
|---|---|---|---|---|---|
| Bill and Melinda Gates Foundation | ☑ | ☐ | ☐ | ☐ | Grant to RFI |
| Jacobs Institute for Molecular Engineering for Medicine (Caltech) | ☑ | ☐ | ☐ | ☐ | Grant to RFI |

#### Section 3. Relevant financial activities outside the submitted work.

Place a check in the appropriate boxes in the table to indicate whether you have financial relationships (regardless of amount of compensation) with entities as described in the instructions. Use one line for each entity; add as many lines as you need by clicking the "Add +" box. You should report relationships that were **present during the 36 months prior to publication**.

Are there any relevant conflicts of interest? ☐ Yes ☑ No

#### Section 4. Intellectual Property -- Patents & Copyrights

Do you have any patents, whether planned, pending or issued, broadly relevant to the work? ☐ Yes ☑ No

ICMJE INTERNATIONAL COMMITTEE of MEDICAL JOURNAL EDITORS

### ICMJE Form for Disclosure of Potential Conflicts of Interest

#### Section 5. Relationships not covered above

Are there other relationships or activities that readers could perceive to have influenced, or that give the appearance of potentially influencing, what you wrote in the submitted work?

☐ Yes, the following relationships/conditions/circumstances are present (explain below):

☑ No other relationships/conditions/circumstances that present a potential conflict of interest

At the time of manuscript acceptance, journals will ask authors to confirm and, if necessary, update their disclosure statements. On occasion, journals may ask authors to disclose further information about reported relationships.

#### Section 6. Disclosure Statement

Based on the above disclosures, this form will automatically generate a disclosure statement, which will appear in the box below.

Mr. Akana reports grants (to RFI) from Bill & Melinda Gates Foundation and the Jacobs Institute for Molecular Engineering for Medicine (Caltech) , during the conduct of the study; .

#### Evaluation and Feedback

Please visit http://www.icmje.org/cgi-bin/feedback to provide feedback on your experience with completing this form.

ICMJE INTERNATIONAL COMMITTEE of MEDICAL JOURNAL EDITORS

### ICMJE Form for Disclosure of Potential Conflicts of Interest

#### Instructions

**The purpose of this form is to provide readers of your manuscript with information about your other interests that could influence how they receive and understand your work. The form is designed to be completed electronically and stored electronically. It contains programming that allows appropriate data display. Each author should submit a separate form and is responsible for the accuracy and completeness of the submitted information. The form is in six parts.**

#### 1. Identifying information.

#### 2. The work under consideration for publication.

This section asks for information about the work that you have submitted for publication. The time frame for this reporting is that of the work itself, from the initial conception and planning to the present. The requested information is about resources that you received, either directly or indirectly (via your institution), to enable you to complete the work. Checking "No" means that you did the work without receiving any financial support from any third party -- that is, the work was supported by funds from the same institution that pays your salary and that institution did not receive third-party funds with which to pay you. If you or your institution received funds from a third party to support the work, such as a government granting agency, charitable foundation or commercial sponsor, check "Yes".

#### 3. Relevant financial activities outside the submitted work.

This section asks about your financial relationships with entities in the bio-medical arena that could be perceived to influence, or that give the appearance of potentially influencing, what you wrote in the submitted work. You should disclose interactions with ANY entity that could be considered broadly relevant to the work. For example, if your article is about testing an epidermal growth factor receptor (EGFR) antagonist in lung cancer, you should report all associations with entities pursuing diagnostic or therapeutic strategies in cancer in general, not just in the area of EGFR or lung cancer.

Report all sources of revenue paid (or promised to be paid) directly to you or your institution on your behalf over the 36 months prior to submission of the work. This should include all monies from sources with relevance to the submitted work, not just monies from the entity that sponsored the research. Please note that your interactions with the work's sponsor that are outside the submitted work should also be listed here. If there is any question, it is usually better to disclose a relationship than not to do so.

For grants you have received for work outside the submitted work, you should disclose support ONLY from entities that could be perceived to be affected financially by the published work, such as drug companies, or foundations supported by entities that could be perceived to have a financial stake in the outcome. Public funding sources, such as government agencies, charitable foundations or academic institutions, need not be disclosed. For example, if a government agency sponsored a study in which you have been involved and drugs were provided by a pharmaceutical company, you need only list the pharmaceutical company.

#### 4. Intellectual Property.

This section asks about patents and copyrights, whether pending, issued, licensed and/or receiving royalties.

#### 5. Relationships not covered above.

Use this section to report other relationships or activities that readers could perceive to have influenced, or that give the appearance of potentially influencing, what you wrote in the submitted work.

##### Definitions.

**Entity:** government agency, foundation, commercial sponsor, academic institution, etc.
**Grant:** A grant from an entity, generally [but not always] paid to your organization
**Personal Fees:** Monies paid to you for services rendered, generally honoraria, royalties, or fees for consulting , lectures, speakers bureaus, expert testimony, employment, or other affiliations
**Non-Financial Support:** Examples include drugs/equipment supplied by the entity, travel paid by the entity, writing assistance, administrative support, etc**.**

**Other:** Anything not covered under the previous three boxes
**Pending:** The patent has been filed but not issued
**Issued:** The patent has been issued by the agency
**Licensed:** The patent has been licensed to an entity, whether earning royalties or not
**Royalties:** Funds are coming in to you or your institution due to your patent

### ICMJE Form for Disclosure of Potential Conflicts of Interest

#### Section 1. Identifying Information

1. Given Name (First Name)
Jacob

2. Surname (Last Name)
Barlow

3. Date
22-November-2020

4. Are you the corresponding author? ☐ Yes ☑ No

Corresponding Author's Name
Rustem F. Ismagilov

5. Manuscript Title
SARS-CoV-2 Viral Load Rises Gradually and to Moderate Levels in Some Humans

6. Manuscript Identifying Number (if you know it)

#### Section 2. The Work Under Consideration for Publication

Did you or your institution **at any time** receive payment or services from a third party (government, commercial, private foundation, etc.) for any aspect of the submitted work (including but not limited to grants, data monitoring board, study design, manuscript preparation, statistical analysis, etc.)?

Are there any relevant conflicts of interest? ☑ Yes ☐ No

If yes, please fill out the appropriate information below. If you have more than one entity press the "ADD" button to add a row. Excess rows can be removed by pressing the "X" button.

| Name of Institution/Company | Grant? | Personal Fees? | Non-Financial Support? | Other? | Comments |
|---|---|---|---|---|---|
| Jacobs Institute for Molecular Engineering for Medicine (Caltech) | ☑ | ☐ | ☐ | ☐ | Grant to RFI |
| Bill & Melinda Gates Foundation | ☑ | ☐ | ☐ | ☐ | Grant to RFI |
| National Institutes of Health Biotechnology Leadership Pre-doctoral Training Program (BLP) fellowships from Caltech's Donna and Benjamin M. Rosen Bioengineering Center (T32GM112592) | ☐ | ☐ | ☐ | ☑ | Fellowship to JB |

#### Section 3. Relevant financial activities outside the submitted work.

Place a check in the appropriate boxes in the table to indicate whether you have financial relationships (regardless of amount of compensation) with entities as described in the instructions. Use one line for each entity; add as many lines as you need by clicking the "Add +" box. You should report relationships that were **present during the 36 months prior to publication**.

Are there any relevant conflicts of interest? ☐ Yes ☑ No

INTERNATIONAL COMMITTEE of MEDICAL JOURNAL EDITORS

### ICMJE Form for Disclosure of Potential Conflicts of Interest

#### Section 4. Intellectual Property -- Patents & Copyrights

Do you have any patents, whether planned, pending or issued, broadly relevant to the work? ☐ Yes ☑ No

#### Section 5. Relationships not covered above

Are there other relationships or activities that readers could perceive to have influenced, or that give the appearance of potentially influencing, what you wrote in the submitted work?

☐ Yes, the following relationships/conditions/circumstances are present (explain below):

☑ No other relationships/conditions/circumstances that present a potential conflict of interest

At the time of manuscript acceptance, journals will ask authors to confirm and, if necessary, update their disclosure statements. On occasion, journals may ask authors to disclose further information about reported relationships.

#### Section 6. Disclosure Statement

Based on the above disclosures, this form will automatically generate a disclosure statement, which will appear in the box below.

Jacob Barlow reports grants (to RFI) from the Jacobs Institute for Molecular Engineering for Medicine (Caltech) and the Bill & Melinda Gates Foundation, and a National Institutes of Health Biotechnology Leadership Pre-doctoral Training Program (BLP) fellowship from Caltech's Donna and Benjamin M. Rosen Bioengineering Center (T32GM112592), during the conduct of the study; .

#### Evaluation and Feedback

Please visit http://www.icmje.org/cgi-bin/feedback to provide feedback on your experience with completing this form.

INTERNATIONAL COMMITTEE of MEDICAL JOURNAL EDITORS

### ICMJE Form for Disclosure of Potential Conflicts of Interest

#### Instructions

**The purpose of this form is to provide readers of your manuscript with information about your other interests that could influence how they receive and understand your work. The form is designed to be completed electronically and stored electronically. It contains programming that allows appropriate data display. Each author should submit a separate form and is responsible for the accuracy and completeness of the submitted information. The form is in six parts.**

#### 1. Identifying information.

#### 2. The work under consideration for publication.

This section asks for information about the work that you have submitted for publication. The time frame for this reporting is that of the work itself, from the initial conception and planning to the present. The requested information is about resources that you received, either directly or indirectly (via your institution), to enable you to complete the work. Checking "No" means that you did the work without receiving any financial support from any third party -- that is, the work was supported by funds from the same institution that pays your salary and that institution did not receive third-party funds with which to pay you. If you or your institution received funds from a third party to support the work, such as a government granting agency, charitable foundation or commercial sponsor, check "Yes".

#### 3. Relevant financial activities outside the submitted work.

This section asks about your financial relationships with entities in the bio-medical arena that could be perceived to influence, or that give the appearance of potentially influencing, what you wrote in the submitted work. You should disclose interactions with ANY entity that could be considered broadly relevant to the work. For example, if your article is about testing an epidermal growth factor receptor (EGFR) antagonist in lung cancer, you should report all associations with entities pursuing diagnostic or therapeutic strategies in cancer in general, not just in the area of EGFR or lung cancer.

Report all sources of revenue paid (or promised to be paid) directly to you or your institution on your behalf over the 36 months prior to submission of the work. This should include all monies from sources with relevance to the submitted work, not just monies from the entity that sponsored the research. Please note that your interactions with the work's sponsor that are outside the submitted work should also be listed here. If there is any question, it is usually better to disclose a relationship than not to do so.

For grants you have received for work outside the submitted work, you should disclose support ONLY from entities that could be perceived to be affected financially by the published work, such as drug companies, or foundations supported by entities that could be perceived to have a financial stake in the outcome. Public funding sources, such as government agencies, charitable foundations or academic institutions, need not be disclosed. For example, if a government agency sponsored a study in which you have been involved and drugs were provided by a pharmaceutical company, you need only list the pharmaceutical company.

#### 4. Intellectual Property.

This section asks about patents and copyrights, whether pending, issued, licensed and/or receiving royalties.

#### 5. Relationships not covered above.

Use this section to report other relationships or activities that readers could perceive to have influenced, or that give the appearance of potentially influencing, what you wrote in the submitted work.

##### Definitions.

**Entity:** government agency, foundation, commercial sponsor, academic institution, etc.
**Grant:** A grant from an entity, generally [but not always] paid to your organization
**Personal Fees:** Monies paid to you for services rendered, generally honoraria, royalties, or fees for consulting , lectures, speakers bureaus, expert testimony, employment, or other affiliations
**Non-Financial Support:** Examples include drugs/equipment supplied by the entity, travel paid by the entity, writing assistance, administrative support, etc**.**

**Other:** Anything not covered under the previous three boxes
**Pending:** The patent has been filed but not issued
**Issued:** The patent has been issued by the agency
**Licensed:** The patent has been licensed to an entity, whether earning royalties or not
**Royalties:** Funds are coming in to you or your institution due to your patent

ICMJE INTERNATIONAL COMMITTEE of MEDICAL JOURNAL EDITORS

### ICMJE Form for Disclosure of Potential Conflicts of Interest

#### Section 1. Identifying Information

1. Given Name (First Name)
Matthew

2. Surname (Last Name)
Cooper

3. Date
20-November-2020

4. Are you the corresponding author? ☐ Yes ☑ No

Corresponding Author's Name
Rustem F. Ismagilov

5. Manuscript Title
SARS-CoV-2 Viral Load Rises Gradually and to Moderate Levels in Some Humans

6. Manuscript Identifying Number (if you know it)
n/a

#### Section 2. The Work Under Consideration for Publication

Did you or your institution **at any time** receive payment or services from a third party (government, commercial, private foundation, etc.) for any aspect of the submitted work (including but not limited to grants, data monitoring board, study design, manuscript preparation, statistical analysis, etc.)?

Are there any relevant conflicts of interest? ☑ Yes ☐ No

If yes, please fill out the appropriate information below. If you have more than one entity press the "ADD" button to add a row. Excess rows can be removed by pressing the "X" button.

| Name of Institution/Company | Grant? | Personal Fees? | Non-Financial Support? | Other? | Comments |
|---|---|---|---|---|---|
| Caltech Graduate Student Fellowship | ☐ | ☐ | ☐ | ☑ | fellowship |
| Bill & Melinda Gates Foundation | ☑ | ☐ | ☐ | ☐ | Grant to RFI |
| Jacobs Institute for Molecular Engineering for Medicine (Caltech) | ☑ | ☐ | ☐ | ☐ | Grant to RFI |

#### Section 3. Relevant financial activities outside the submitted work.

Place a check in the appropriate boxes in the table to indicate whether you have financial relationships (regardless of amount of compensation) with entities as described in the instructions. Use one line for each entity; add as many lines as you need by clicking the "Add +" box. You should report relationships that were **present during the 36 months prior to publication**.

Are there any relevant conflicts of interest? ☐ Yes ☑ No

#### Section 4. Intellectual Property -- Patents & Copyrights

Do you have any patents, whether planned, pending or issued, broadly relevant to the work? ☐ Yes ☑ No

ICMJE INTERNATIONAL COMMITTEE of MEDICAL JOURNAL EDITORS

### ICMJE Form for Disclosure of Potential Conflicts of Interest

#### Section 5. Relationships not covered above

Are there other relationships or activities that readers could perceive to have influenced, or that give the appearance of potentially influencing, what you wrote in the submitted work?

☐ Yes, the following relationships/conditions/circumstances are present (explain below):

☑ No other relationships/conditions/circumstances that present a potential conflict of interest

At the time of manuscript acceptance, journals will ask authors to confirm and, if necessary, update their disclosure statements. On occasion, journals may ask authors to disclose further information about reported relationships.

#### Section 6. Disclosure Statement

Based on the above disclosures, this form will automatically generate a disclosure statement, which will appear in the box below.

Mr. Cooper reports funding from a Caltech Graduate Student Fellowship, and grants (to RFI) from Bill & Melinda Gates Foundation and the Jacobs Institute for Molecular Engineering for Medicine (Caltech), during the conduct of the study; .

#### Evaluation and Feedback

Please visit http://www.icmje.org/cgi-bin/feedback to provide feedback on your experience with completing this form.

ICMJE INTERNATIONAL COMMITTEE of MEDICAL JOURNAL EDITORS

### ICMJE Form for Disclosure of Potential Conflicts of Interest

#### Instructions

**The purpose of this form is to provide readers of your manuscript with information about your other interests that could influence how they receive and understand your work. The form is designed to be completed electronically and stored electronically. It contains programming that allows appropriate data display. Each author should submit a separate form and is responsible for the accuracy and completeness of the submitted information. The form is in six parts.**

#### 1. Identifying information.

#### 2. The work under consideration for publication.

This section asks for information about the work that you have submitted for publication. The time frame for this reporting is that of the work itself, from the initial conception and planning to the present. The requested information is about resources that you received, either directly or indirectly (via your institution), to enable you to complete the work. Checking "No" means that you did the work without receiving any financial support from any third party -- that is, the work was supported by funds from the same institution that pays your salary and that institution did not receive third-party funds with which to pay you. If you or your institution received funds from a third party to support the work, such as a government granting agency, charitable foundation or commercial sponsor, check "Yes".

#### 3. Relevant financial activities outside the submitted work.

This section asks about your financial relationships with entities in the bio-medical arena that could be perceived to influence, or that give the appearance of potentially influencing, what you wrote in the submitted work. You should disclose interactions with ANY entity that could be considered broadly relevant to the work. For example, if your article is about testing an epidermal growth factor receptor (EGFR) antagonist in lung cancer, you should report all associations with entities pursuing diagnostic or therapeutic strategies in cancer in general, not just in the area of EGFR or lung cancer.

Report all sources of revenue paid (or promised to be paid) directly to you or your institution on your behalf over the 36 months prior to submission of the work. This should include all monies from sources with relevance to the submitted work, not just monies from the entity that sponsored the research. Please note that your interactions with the work's sponsor that are outside the submitted work should also be listed here. If there is any question, it is usually better to disclose a relationship than not to do so.

For grants you have received for work outside the submitted work, you should disclose support ONLY from entities that could be perceived to be affected financially by the published work, such as drug companies, or foundations supported by entities that could be perceived to have a financial stake in the outcome. Public funding sources, such as government agencies, charitable foundations or academic institutions, need not be disclosed. For example, if a government agency sponsored a study in which you have been involved and drugs were provided by a pharmaceutical company, you need only list the pharmaceutical company.

#### 4. Intellectual Property.

This section asks about patents and copyrights, whether pending, issued, licensed and/or receiving royalties.

#### 5. Relationships not covered above.

Use this section to report other relationships or activities that readers could perceive to have influenced, or that give the appearance of potentially influencing, what you wrote in the submitted work.

##### Definitions.

**Entity:** government agency, foundation, commercial sponsor, academic institution, etc.
**Grant:** A grant from an entity, generally [but not always] paid to your organization
**Personal Fees:** Monies paid to you for services rendered, generally honoraria, royalties, or fees for consulting , lectures, speakers bureaus, expert testimony, employment, or other affiliations
**Non-Financial Support:** Examples include drugs/equipment supplied by the entity, travel paid by the entity, writing assistance, administrative support, etc.

**Other:** Anything not covered under the previous three boxes
**Pending:** The patent has been filed but not issued
**Issued:** The patent has been issued by the agency
**Licensed:** The patent has been licensed to an entity, whether earning royalties or not
**Royalties:** Funds are coming in to you or your institution due to your patent

### ICMJE Form for Disclosure of Potential Conflicts of Interest

#### Section 1. Identifying Information

1. Given Name (First Name)
Matthew

2. Surname (Last Name)
Feaster

3. Date
22-November-2020

4. Are you the corresponding author? ☐ Yes ☑ No

Corresponding Author's Name
Rustem F. Ismagilov

5. Manuscript Title
SARS-CoV-2 Viral Load Rises Gradually and to Moderate Levels in Some Humans

6. Manuscript Identifying Number (if you know it)

#### Section 2. The Work Under Consideration for Publication

Did you or your institution **at any time** receive payment or services from a third party (government, commercial, private foundation, etc.) for any aspect of the submitted work (including but not limited to grants, data monitoring board, study design, manuscript preparation, statistical analysis, etc.)?

Are there any relevant conflicts of interest? ☐ Yes ☑ No

#### Section 3. Relevant financial activities outside the submitted work.

Place a check in the appropriate boxes in the table to indicate whether you have financial relationships (regardless of amount of compensation) with entities as described in the instructions. Use one line for each entity; add as many lines as you need by clicking the "Add +" box. You should report relationships that were **present during the 36 months prior to publication**.

Are there any relevant conflicts of interest? ☐ Yes ☑ No

#### Section 4. Intellectual Property -- Patents & Copyrights

Do you have any patents, whether planned, pending or issued, broadly relevant to the work? ☐ Yes ☑ No

### ICMJE Form for Disclosure of Potential Conflicts of Interest

#### Section 5. Relationships not covered above

Are there other relationships or activities that readers could perceive to have influenced, or that give the appearance of potentially influencing, what you wrote in the submitted work?

☐ Yes, the following relationships/conditions/circumstances are present (explain below):

☑ No other relationships/conditions/circumstances that present a potential conflict of interest

At the time of manuscript acceptance, journals will ask authors to confirm and, if necessary, update their disclosure statements. On occasion, journals may ask authors to disclose further information about reported relationships.

#### Section 6. Disclosure Statement

Based on the above disclosures, this form will automatically generate a disclosure statement, which will appear in the box below.

Dr. Feaster has nothing to disclose.

#### Evaluation and Feedback

Please visit http://www.icmje.org/cgi-bin/feedback to provide feedback on your experience with completing this form.

ICMJE INTERNATIONAL COMMITTEE of MEDICAL JOURNAL EDITORS

### ICMJE Form for Disclosure of Potential Conflicts of Interest

#### Instructions

**The purpose of this form is to provide readers of your manuscript with information about your other interests that could influence how they receive and understand your work. The form is designed to be completed electronically and stored electronically. It contains programming that allows appropriate data display. Each author should submit a separate form and is responsible for the accuracy and completeness of the submitted information. The form is in six parts.**

#### 1. Identifying information.

#### 2. The work under consideration for publication.

This section asks for information about the work that you have submitted for publication. The time frame for this reporting is that of the work itself, from the initial conception and planning to the present. The requested information is about resources that you received, either directly or indirectly (via your institution), to enable you to complete the work. Checking "No" means that you did the work without receiving any financial support from any third party -- that is, the work was supported by funds from the same institution that pays your salary and that institution did not receive third-party funds with which to pay you. If you or your institution received funds from a third party to support the work, such as a government granting agency, charitable foundation or commercial sponsor, check "Yes".

#### 3. Relevant financial activities outside the submitted work.

This section asks about your financial relationships with entities in the bio-medical arena that could be perceived to influence, or that give the appearance of potentially influencing, what you wrote in the submitted work. You should disclose interactions with ANY entity that could be considered broadly relevant to the work. For example, if your article is about testing an epidermal growth factor receptor (EGFR) antagonist in lung cancer, you should report all associations with entities pursuing diagnostic or therapeutic strategies in cancer in general, not just in the area of EGFR or lung cancer.

Report all sources of revenue paid (or promised to be paid) directly to you or your institution on your behalf over the 36 months prior to submission of the work. This should include all monies from sources with relevance to the submitted work, not just monies from the entity that sponsored the research. Please note that your interactions with the work's sponsor that are outside the submitted work should also be listed here. If there is any question, it is usually better to disclose a relationship than not to do so.

For grants you have received for work outside the submitted work, you should disclose support ONLY from entities that could be perceived to be affected financially by the published work, such as drug companies, or foundations supported by entities that could be perceived to have a financial stake in the outcome. Public funding sources, such as government agencies, charitable foundations or academic institutions, need not be disclosed. For example, if a government agency sponsored a study in which you have been involved and drugs were provided by a pharmaceutical company, you need only list the pharmaceutical company.

#### 4. Intellectual Property.

This section asks about patents and copyrights, whether pending, issued, licensed and/or receiving royalties.

#### 5. Relationships not covered above.

Use this section to report other relationships or activities that readers could perceive to have influenced, or that give the appearance of potentially influencing, what you wrote in the submitted work.

##### Definitions.

**Entity:** government agency, foundation, commercial sponsor, academic institution, etc.
**Grant:** A grant from an entity, generally [but not always] paid to your organization
**Personal Fees:** Monies paid to you for services rendered, generally honoraria, royalties, or fees for consulting , lectures, speakers bureaus, expert testimony, employment, or other affiliations
**Non-Financial Support:** Examples include drugs/equipment supplied by the entity, travel paid by the entity, writing assistance, administrative support, etc**.**

**Other:** Anything not covered under the previous three boxes
**Pending:** The patent has been filed but not issued
**Issued:** The patent has been issued by the agency
**Licensed:** The patent has been licensed to an entity, whether earning royalties or not
**Royalties:** Funds are coming in to you or your institution due to your patent

### ICMJE Form for Disclosure of Potential Conflicts of Interest

#### Section 1. Identifying Information

1. Given Name (First Name)
Ying-Ying

2. Surname (Last Name)
Goh

3. Date
22-November-2020

4. Are you the corresponding author? ☐ Yes ☑ No

Corresponding Author's Name
Rustem F. Ismagilov

5. Manuscript Title
SARS-CoV-2 Viral Load Rises Gradually and to Moderate Levels in Some Humans

6. Manuscript Identifying Number (if you know it)

#### Section 2. The Work Under Consideration for Publication

Did you or your institution **at any time** receive payment or services from a third party (government, commercial, private foundation, etc.) for any aspect of the submitted work (including but not limited to grants, data monitoring board, study design, manuscript preparation, statistical analysis, etc.)?

Are there any relevant conflicts of interest? ☐ Yes ☑ No

#### Section 3. Relevant financial activities outside the submitted work.

Place a check in the appropriate boxes in the table to indicate whether you have financial relationships (regardless of amount of compensation) with entities as described in the instructions. Use one line for each entity; add as many lines as you need by clicking the "Add +" box. You should report relationships that were **present during the 36 months prior to publication**.

Are there any relevant conflicts of interest? ☐ Yes ☑ No

#### Section 4. Intellectual Property -- Patents & Copyrights

Do you have any patents, whether planned, pending or issued, broadly relevant to the work? ☐ Yes ☑ No

### ICMJE Form for Disclosure of Potential Conflicts of Interest

#### Section 5. Relationships not covered above

Are there other relationships or activities that readers could perceive to have influenced, or that give the appearance of potentially influencing, what you wrote in the submitted work?

☐ Yes, the following relationships/conditions/circumstances are present (explain below):

☑ No other relationships/conditions/circumstances that present a potential conflict of interest

At the time of manuscript acceptance, journals will ask authors to confirm and, if necessary, update their disclosure statements. On occasion, journals may ask authors to disclose further information about reported relationships.

#### Section 6. Disclosure Statement

Based on the above disclosures, this form will automatically generate a disclosure statement, which will appear in the box below.

Dr. Goh has nothing to disclose.

#### Evaluation and Feedback

Please visit http://www.icmje.org/cgi-bin/feedback to provide feedback on your experience with completing this form.

### ICMJE Form for Disclosure of Potential Conflicts of Interest

#### Instructions

**The purpose of this form is to provide readers of your manuscript with information about your other interests that could influence how they receive and understand your work. The form is designed to be completed electronically and stored electronically. It contains programming that allows appropriate data display. Each author should submit a separate form and is responsible for the accuracy and completeness of the submitted information. The form is in six parts.**

#### 1. Identifying information.

#### 2. The work under consideration for publication.

This section asks for information about the work that you have submitted for publication. The time frame for this reporting is that of the work itself, from the initial conception and planning to the present. The requested information is about resources that you received, either directly or indirectly (via your institution), to enable you to complete the work. Checking "No" means that you did the work without receiving any financial support from any third party -- that is, the work was supported by funds from the same institution that pays your salary and that institution did not receive third-party funds with which to pay you. If you or your institution received funds from a third party to support the work, such as a government granting agency, charitable foundation or commercial sponsor, check "Yes".

#### 3. Relevant financial activities outside the submitted work.

This section asks about your financial relationships with entities in the bio-medical arena that could be perceived to influence, or that give the appearance of potentially influencing, what you wrote in the submitted work. You should disclose interactions with ANY entity that could be considered broadly relevant to the work. For example, if your article is about testing an epidermal growth factor receptor (EGFR) antagonist in lung cancer, you should report all associations with entities pursuing diagnostic or therapeutic strategies in cancer in general, not just in the area of EGFR or lung cancer.

Report all sources of revenue paid (or promised to be paid) directly to you or your institution on your behalf over the 36 months prior to submission of the work. This should include all monies from sources with relevance to the submitted work, not just monies from the entity that sponsored the research. Please note that your interactions with the work's sponsor that are outside the submitted work should also be listed here. If there is any question, it is usually better to disclose a relationship than not to do so.

For grants you have received for work outside the submitted work, you should disclose support ONLY from entities that could be perceived to be affected financially by the published work, such as drug companies, or foundations supported by entities that could be perceived to have a financial stake in the outcome. Public funding sources, such as government agencies, charitable foundations or academic institutions, need not be disclosed. For example, if a government agency sponsored a study in which you have been involved and drugs were provided by a pharmaceutical company, you need only list the pharmaceutical company.

#### 4. Intellectual Property.

This section asks about patents and copyrights, whether pending, issued, licensed and/or receiving royalties.

#### 5. Relationships not covered above.

Use this section to report other relationships or activities that readers could perceive to have influenced, or that give the appearance of potentially influencing, what you wrote in the submitted work.

##### Definitions.

**Entity:** government agency, foundation, commercial sponsor, academic institution, etc.
**Grant:** A grant from an entity, generally [but not always] paid to your organization
**Personal Fees:** Monies paid to you for services rendered, generally honoraria, royalties, or fees for consulting , lectures, speakers bureaus, expert testimony, employment, or other affiliations
**Non-Financial Support:** Examples include drugs/equipment supplied by the entity, travel paid by the entity, writing assistance, administrative support, etc**.**

**Other:** Anything not covered under the previous three boxes
**Pending:** The patent has been filed but not issued
**Issued:** The patent has been issued by the agency
**Licensed:** The patent has been licensed to an entity, whether earning royalties or not
**Royalties:** Funds are coming in to you or your institution due to your patent

### ICMJE Form for Disclosure of Potential Conflicts of Interest

#### Section 1. Identifying Information

1. Given Name (First Name): Rustem
2. Surname (Last Name): Ismagilov
3. Date: 17-November-2020

4. Are you the corresponding author? ☑ Yes ☐ No

5. Manuscript Title
SARS-CoV-2 Viral Load Rises Gradually and to Moderate Levels in Some Humans

6. Manuscript Identifying Number (if you know it)
n/a

#### Section 2. The Work Under Consideration for Publication

Did you or your institution **at any time** receive payment or services from a third party (government, commercial, private foundation, etc.) for any aspect of the submitted work (including but not limited to grants, data monitoring board, study design, manuscript preparation, statistical analysis, etc.)?

Are there any relevant conflicts of interest? ☑ Yes ☐ No

If yes, please fill out the appropriate information below. If you have more than one entity press the "ADD" button to add a row. Excess rows can be removed by pressing the "X" button.

| Name of Institution/Company | Grant? | Personal Fees? | Non-Financial Support? | Other? | Comments |
|---|---|---|---|---|---|
| Bill & Melinda Gates Foundation | ☑ | ☐ | ☐ | ☐ | |
| Jacobs Institute for Molecular Engineering for Medicine (Caltech) | ☑ | ☐ | ☐ | ☐ | |

#### Section 3. Relevant financial activities outside the submitted work.

Place a check in the appropriate boxes in the table to indicate whether you have financial relationships (regardless of amount of compensation) with entities as described in the instructions. Use one line for each entity; add as many lines as you need by clicking the "Add +" box. You should report relationships that were **present during the 36 months prior to publication**.

Are there any relevant conflicts of interest? ☑ Yes ☐ No

If yes, please fill out the appropriate information below.

ICMJE INTERNATIONAL COMMITTEE of MEDICAL JOURNAL EDITORS

### ICMJE Form for Disclosure of Potential Conflicts of Interest

| Name of Entity | Grant? | Personal Fees? | Non-Financial Support? | Other? | Comments |
|---|---|---|---|---|---|
| Talis Biomedical Corp. | ☐ | ☑ | ☐ | ☑ | co-Founder, Consultant, Board of Directors |

#### Section 4. Intellectual Property -- Patents & Copyrights

Do you have any patents, whether planned, pending or issued, broadly relevant to the work? ☑ Yes ☐ No

If yes, please fill out the appropriate information below. If you have more than one entity press the "ADD" button to add a row. Excess rows can be removed by pressing the "X" button.

| Patent? | Pending? | Issued? | Licensed? | Royalties? | Licensee? | Comments |
|---|---|---|---|---|---|---|
| US Patent 8,304,193. Method for conducting an autocatalytic reaction in plugs in a microfluidic system | ☐ | ☑ | ☑ | ☑ | Bio-Rad Laboratories Inc. | |

#### Section 5. Relationships not covered above

Are there other relationships or activities that readers could perceive to have influenced, or that give the appearance of potentially influencing, what you wrote in the submitted work?

☐ Yes, the following relationships/conditions/circumstances are present (explain below):

☑ No other relationships/conditions/circumstances that present a potential conflict of interest

At the time of manuscript acceptance, journals will ask authors to confirm and, if necessary, update their disclosure statements. On occasion, journals may ask authors to disclose further information about reported relationships.

### ICMJE Form for Disclosure of Potential Conflicts of Interest

#### Section 6. Disclosure Statement

Based on the above disclosures, this form will automatically generate a disclosure statement, which will appear in the box below.

Dr. Ismagilov reports grants from the Bill & Melinda Gates Foundation and the Jacobs Institute for Molecular Engineering for Medicine (Caltech), during the conduct of the study. Dr. Ismagilov is a co-founder, consultant, and a Director and has stock ownership and received personal fees from Talis Biomedical Corp., outside the submitted work. In addition, Dr. Ismagilov has a series of patents including US Patent 8,304,193, "Method for conducting an autocatalytic reaction in plugs in a microfluidic system" licensed with royalties paid to Bio-Rad Laboratories Inc. in the context of ddPCR.

#### Evaluation and Feedback

Please visit http://www.icmje.org/cgi-bin/feedback to provide feedback on your experience with completing this form.

INTERNATIONAL COMMITTEE of MEDICAL JOURNAL EDITORS

### ICMJE Form for Disclosure of Potential Conflicts of Interest

#### Instructions

**The purpose of this form is to provide readers of your manuscript with information about your other interests that could influence how they receive and understand your work. The form is designed to be completed electronically and stored electronically. It contains programming that allows appropriate data display. Each author should submit a separate form and is responsible for the accuracy and completeness of the submitted information. The form is in six parts.**

#### 1. Identifying information.

#### 2. The work under consideration for publication.

This section asks for information about the work that you have submitted for publication. The time frame for this reporting is that of the work itself, from the initial conception and planning to the present. The requested information is about resources that you received, either directly or indirectly (via your institution), to enable you to complete the work. Checking "No" means that you did the work without receiving any financial support from any third party -- that is, the work was supported by funds from the same institution that pays your salary and that institution did not receive third-party funds with which to pay you. If you or your institution received funds from a third party to support the work, such as a government granting agency, charitable foundation or commercial sponsor, check "Yes".

#### 3. Relevant financial activities outside the submitted work.

This section asks about your financial relationships with entities in the bio-medical arena that could be perceived to influence, or that give the appearance of potentially influencing, what you wrote in the submitted work. You should disclose interactions with ANY entity that could be considered broadly relevant to the work. For example, if your article is about testing an epidermal growth factor receptor (EGFR) antagonist in lung cancer, you should report all associations with entities pursuing diagnostic or therapeutic strategies in cancer in general, not just in the area of EGFR or lung cancer.

Report all sources of revenue paid (or promised to be paid) directly to you or your institution on your behalf over the 36 months prior to submission of the work. This should include all monies from sources with relevance to the submitted work, not just monies from the entity that sponsored the research. Please note that your interactions with the work's sponsor that are outside the submitted work should also be listed here. If there is any question, it is usually better to disclose a relationship than not to do so.

For grants you have received for work outside the submitted work, you should disclose support ONLY from entities that could be perceived to be affected financially by the published work, such as drug companies, or foundations supported by entities that could be perceived to have a financial stake in the outcome. Public funding sources, such as government agencies, charitable foundations or academic institutions, need not be disclosed. For example, if a government agency sponsored a study in which you have been involved and drugs were provided by a pharmaceutical company, you need only list the pharmaceutical company.

#### 4. Intellectual Property.

This section asks about patents and copyrights, whether pending, issued, licensed and/or receiving royalties.

#### 5. Relationships not covered above.

Use this section to report other relationships or activities that readers could perceive to have influenced, or that give the appearance of potentially influencing, what you wrote in the submitted work.

##### Definitions.

**Entity:** government agency, foundation, commercial sponsor, academic institution, etc.
**Grant:** A grant from an entity, generally [but not always] paid to your organization
**Personal Fees:** Monies paid to you for services rendered, generally honoraria, royalties, or fees for consulting , lectures, speakers bureaus, expert testimony, employment, or other affiliations
**Non-Financial Support:** Examples include drugs/equipment supplied by the entity, travel paid by the entity, writing assistance, administrative support, etc**.**

**Other:** Anything not covered under the previous three boxes
**Pending:** The patent has been filed but not issued
**Issued:** The patent has been issued by the agency
**Licensed:** The patent has been licensed to an entity, whether earning royalties or not
**Royalties:** Funds are coming in to you or your institution due to your patent

### ICMJE Form for Disclosure of Potential Conflicts of Interest

#### Section 1. Identifying Information

1. Given Name (First Name)
Jenny

2. Surname (Last Name)
Ji

3. Date
20-November-2020

4. Are you the corresponding author? ☐ Yes ☑ No

Corresponding Author's Name
Rustem F. Ismagilov

5. Manuscript Title
SARS-CoV-2 Viral Load Rises Gradually and to Moderate Levels in Some Humans

6. Manuscript Identifying Number (if you know it)

#### Section 2. The Work Under Consideration for Publication

Did you or your institution **at any time** receive payment or services from a third party (government, commercial, private foundation, etc.) for any aspect of the submitted work (including but not limited to grants, data monitoring board, study design, manuscript preparation, statistical analysis, etc.)?

Are there any relevant conflicts of interest? ☐ Yes ☑ No

#### Section 3. Relevant financial activities outside the submitted work.

Place a check in the appropriate boxes in the table to indicate whether you have financial relationships (regardless of amount of compensation) with entities as described in the instructions. Use one line for each entity; add as many lines as you need by clicking the "Add +" box. You should report relationships that were **present during the 36 months prior to publication**.

Are there any relevant conflicts of interest? ☐ Yes ☑ No

#### Section 4. Intellectual Property -- Patents & Copyrights

Do you have any patents, whether planned, pending or issued, broadly relevant to the work? ☐ Yes ☑ No

### ICMJE Form for Disclosure of Potential Conflicts of Interest

#### Section 5. Relationships not covered above

Are there other relationships or activities that readers could perceive to have influenced, or that give the appearance of potentially influencing, what you wrote in the submitted work?

☐ Yes, the following relationships/conditions/circumstances are present (explain below):

☑ No other relationships/conditions/circumstances that present a potential conflict of interest

At the time of manuscript acceptance, journals will ask authors to confirm and, if necessary, update their disclosure statements. On occasion, journals may ask authors to disclose further information about reported relationships.

#### Section 6. Disclosure Statement

Based on the above disclosures, this form will automatically generate a disclosure statement, which will appear in the box below.

Jenny Ji has nothing to disclose.

#### Evaluation and Feedback

Please visit http://www.icmje.org/cgi-bin/feedback to provide feedback on your experience with completing this form.

### ICMJE Form for Disclosure of Potential Conflicts of Interest

#### Instructions

**The purpose of this form is to provide readers of your manuscript with information about your other interests that could influence how they receive and understand your work. The form is designed to be completed electronically and stored electronically. It contains programming that allows appropriate data display. Each author should submit a separate form and is responsible for the accuracy and completeness of the submitted information. The form is in six parts.**

#### 1. Identifying information.

#### 2. The work under consideration for publication.

This section asks for information about the work that you have submitted for publication. The time frame for this reporting is that of the work itself, from the initial conception and planning to the present. The requested information is about resources that you received, either directly or indirectly (via your institution), to enable you to complete the work. Checking "No" means that you did the work without receiving any financial support from any third party -- that is, the work was supported by funds from the same institution that pays your salary and that institution did not receive third-party funds with which to pay you. If you or your institution received funds from a third party to support the work, such as a government granting agency, charitable foundation or commercial sponsor, check "Yes".

#### 3. Relevant financial activities outside the submitted work.

This section asks about your financial relationships with entities in the bio-medical arena that could be perceived to influence, or that give the appearance of potentially influencing, what you wrote in the submitted work. You should disclose interactions with ANY entity that could be considered broadly relevant to the work. For example, if your article is about testing an epidermal growth factor receptor (EGFR) antagonist in lung cancer, you should report all associations with entities pursuing diagnostic or therapeutic strategies in cancer in general, not just in the area of EGFR or lung cancer.

Report all sources of revenue paid (or promised to be paid) directly to you or your institution on your behalf over the 36 months prior to submission of the work. This should include all monies from sources with relevance to the submitted work, not just monies from the entity that sponsored the research. Please note that your interactions with the work's sponsor that are outside the submitted work should also be listed here. If there is any question, it is usually better to disclose a relationship than not to do so.

For grants you have received for work outside the submitted work, you should disclose support ONLY from entities that could be perceived to be affected financially by the published work, such as drug companies, or foundations supported by entities that could be perceived to have a financial stake in the outcome. Public funding sources, such as government agencies, charitable foundations or academic institutions, need not be disclosed. For example, if a government agency sponsored a study in which you have been involved and drugs were provided by a pharmaceutical company, you need only list the pharmaceutical company.

#### 4. Intellectual Property.

This section asks about patents and copyrights, whether pending, issued, licensed and/or receiving royalties.

#### 5. Relationships not covered above.

Use this section to report other relationships or activities that readers could perceive to have influenced, or that give the appearance of potentially influencing, what you wrote in the submitted work.

##### Definitions.

**Entity:** government agency, foundation, commercial sponsor, academic institution, etc.
**Grant:** A grant from an entity, generally [but not always] paid to your organization
**Personal Fees:** Monies paid to you for services rendered, generally honoraria, royalties, or fees for consulting , lectures, speakers bureaus, expert testimony, employment, or other affiliations
**Non-Financial Support:** Examples include drugs/equipment supplied by the entity, travel paid by the entity, writing assistance, administrative support, etc**.**

**Other:** Anything not covered under the previous three boxes
**Pending:** The patent has been filed but not issued
**Issued:** The patent has been issued by the agency
**Licensed:** The patent has been licensed to an entity, whether earning royalties or not
**Royalties:** Funds are coming in to you or your institution due to your patent

### ICMJE Form for Disclosure of Potential Conflicts of Interest

#### Section 1. Identifying Information

1. Given Name (First Name)
Michael

2. Surname (Last Name)
Porter

3. Date
20-November-2020

4. Are you the corresponding author? ☐ Yes ☑ No

Corresponding Author's Name
Rustem F. Ismagilov

5. Manuscript Title
SARS-CoV-2 Viral Load Rises Gradually and to Moderate Levels in Some Humans

6. Manuscript Identifying Number (if you know it)

#### Section 2. The Work Under Consideration for Publication

Did you or your institution **at any time** receive payment or services from a third party (government, commercial, private foundation, etc.) for any aspect of the submitted work (including but not limited to grants, data monitoring board, study design, manuscript preparation, statistical analysis, etc.)?

Are there any relevant conflicts of interest? ☑ Yes ☐ No

If yes, please fill out the appropriate information below. If you have more than one entity press the "ADD" button to add a row. Excess rows can be removed by pressing the "X" button.

| Name of Institution/Company | Grant? | Personal Fees? | Non-Financial Support? | Other? | Comments |
|---|---|---|---|---|---|
| Jacobs Institute for Molecular Engineering for Medicine (Caltech) | ☑ | ☐ | ☐ | ☐ | Grant to RFI |
| Bill & Melinda Gates Foundation | ☑ | ☐ | ☐ | ☐ | Grant to RFI |
| National Institutes of Health Biotechnology Leadership Pre-doctoral Training Program (BLP) fellowships from Caltech's Donna and Benjamin M. Rosen Bioengineering Center (T32GM112592) | ☐ | ☐ | ☐ | ☑ | Fellowship to MP |

#### Section 3. Relevant financial activities outside the submitted work.

Place a check in the appropriate boxes in the table to indicate whether you have financial relationships (regardless of amount of compensation) with entities as described in the instructions. Use one line for each entity; add as many lines as you need by clicking the "Add +" box. You should report relationships that were **present during the 36 months prior to publication**.

Are there any relevant conflicts of interest? ☐ Yes ☑ No

INTERNATIONAL COMMITTEE of MEDICAL JOURNAL EDITORS

### ICMJE Form for Disclosure of Potential Conflicts of Interest

#### Section 4. Intellectual Property -- Patents & Copyrights

Do you have any patents, whether planned, pending or issued, broadly relevant to the work? ☐ Yes ☑ No

#### Section 5. Relationships not covered above

Are there other relationships or activities that readers could perceive to have influenced, or that give the appearance of potentially influencing, what you wrote in the submitted work?

☐ Yes, the following relationships/conditions/circumstances are present (explain below):

☑ No other relationships/conditions/circumstances that present a potential conflict of interest

At the time of manuscript acceptance, journals will ask authors to confirm and, if necessary, update their disclosure statements. On occasion, journals may ask authors to disclose further information about reported relationships.

#### Section 6. Disclosure Statement

Based on the above disclosures, this form will automatically generate a disclosure statement, which will appear in the box below.

Mr. Porter reports grants (to RFI) from the Jacobs Institute for Molecular Engineering for Medicine (Caltech) and the Bill & Melinda Gates Foundation, and a National Institutes of Health Biotechnology Leadership Pre-doctoral Training Program (BLP) fellowship from Caltech's Donna and Benjamin M. Rosen Bioengineering Center (T32GM112592), during the conduct of the study; .

#### Evaluation and Feedback

Please visit http://www.icmje.org/cgi-bin/feedback to provide feedback on your experience with completing this form.

### ICMJE Form for Disclosure of Potential Conflicts of Interest

#### Instructions

**The purpose of this form is to provide readers of your manuscript with information about your other interests that could influence how they receive and understand your work. The form is designed to be completed electronically and stored electronically. It contains programming that allows appropriate data display. Each author should submit a separate form and is responsible for the accuracy and completeness of the submitted information. The form is in six parts.**

#### 1. Identifying information.

#### 2. The work under consideration for publication.

This section asks for information about the work that you have submitted for publication. The time frame for this reporting is that of the work itself, from the initial conception and planning to the present. The requested information is about resources that you received, either directly or indirectly (via your institution), to enable you to complete the work. Checking "No" means that you did the work without receiving any financial support from any third party -- that is, the work was supported by funds from the same institution that pays your salary and that institution did not receive third-party funds with which to pay you. If you or your institution received funds from a third party to support the work, such as a government granting agency, charitable foundation or commercial sponsor, check "Yes".

#### 3. Relevant financial activities outside the submitted work.

This section asks about your financial relationships with entities in the bio-medical arena that could be perceived to influence, or that give the appearance of potentially influencing, what you wrote in the submitted work. You should disclose interactions with ANY entity that could be considered broadly relevant to the work. For example, if your article is about testing an epidermal growth factor receptor (EGFR) antagonist in lung cancer, you should report all associations with entities pursuing diagnostic or therapeutic strategies in cancer in general, not just in the area of EGFR or lung cancer.

Report all sources of revenue paid (or promised to be paid) directly to you or your institution on your behalf over the 36 months prior to submission of the work. This should include all monies from sources with relevance to the submitted work, not just monies from the entity that sponsored the research. Please note that your interactions with the work's sponsor that are outside the submitted work should also be listed here. If there is any question, it is usually better to disclose a relationship than not to do so.

For grants you have received for work outside the submitted work, you should disclose support ONLY from entities that could be perceived to be affected financially by the published work, such as drug companies, or foundations supported by entities that could be perceived to have a financial stake in the outcome. Public funding sources, such as government agencies, charitable foundations or academic institutions, need not be disclosed. For example, if a government agency sponsored a study in which you have been involved and drugs were provided by a pharmaceutical company, you need only list the pharmaceutical company.

#### 4. Intellectual Property.

This section asks about patents and copyrights, whether pending, issued, licensed and/or receiving royalties.

#### 5. Relationships not covered above.

Use this section to report other relationships or activities that readers could perceive to have influenced, or that give the appearance of potentially influencing, what you wrote in the submitted work.

##### Definitions.

**Entity:** government agency, foundation, commercial sponsor, academic institution, etc.
**Grant:** A grant from an entity, generally [but not always] paid to your organization
**Personal Fees:** Monies paid to you for services rendered, generally honoraria, royalties, or fees for consulting , lectures, speakers bureaus, expert testimony, employment, or other affiliations
**Non-Financial Support:** Examples include drugs/equipment supplied by the entity, travel paid by the entity, writing assistance, administrative support, etc**.**

**Other:** Anything not covered under the previous three boxes
**Pending:** The patent has been filed but not issued
**Issued:** The patent has been issued by the agency
**Licensed:** The patent has been licensed to an entity, whether earning royalties or not
**Royalties:** Funds are coming in to you or your institution due to your patent

ICMJE INTERNATIONAL COMMITTEE of MEDICAL JOURNAL EDITORS

### ICMJE Form for Disclosure of Potential Conflicts of Interest

#### Section 1. Identifying Information

1. Given Name (First Name)
Jessica

2. Surname (Last Name)
Reyes

3. Date
22-November-2020

4. Are you the corresponding author? ☐ Yes ☑ No

Corresponding Author's Name
Rustem F. Ismagilov

5. Manuscript Title
SARS-CoV-2 Viral Load Rises Gradually and to Moderate Levels in Some Humans

6. Manuscript Identifying Number (if you know it)

#### Section 2. The Work Under Consideration for Publication

Did you or your institution **at any time** receive payment or services from a third party (government, commercial, private foundation, etc.) for any aspect of the submitted work (including but not limited to grants, data monitoring board, study design, manuscript preparation, statistical analysis, etc.)?

Are there any relevant conflicts of interest? ☑ Yes ☐ No

If yes, please fill out the appropriate information below. If you have more than one entity press the "ADD" button to add a row. Excess rows can be removed by pressing the "X" button.

| Name of Institution/Company | Grant? | Personal Fees? | Non-Financial Support? | Other? | Comments |
|---|---|---|---|---|---|
| Bill & Melinda Gates Foundation | ☑ | ☐ | ☐ | ☐ | Grant to RFI |
| Jacobs Institute for Molecular Engineering for Medicine (Caltech) | ☑ | ☐ | ☐ | ☐ | Grant to RFI |

#### Section 3. Relevant financial activities outside the submitted work.

Place a check in the appropriate boxes in the table to indicate whether you have financial relationships (regardless of amount of compensation) with entities as described in the instructions. Use one line for each entity; add as many lines as you need by clicking the "Add +" box. You should report relationships that were **present during the 36 months prior to publication**.

Are there any relevant conflicts of interest? ☐ Yes ☑ No

#### Section 4. Intellectual Property -- Patents & Copyrights

Do you have any patents, whether planned, pending or issued, broadly relevant to the work? ☐ Yes ☑ No

INTERNATIONAL COMMITTEE of MEDICAL JOURNAL EDITORS

### ICMJE Form for Disclosure of Potential Conflicts of Interest

#### Section 5. Relationships not covered above

Are there other relationships or activities that readers could perceive to have influenced, or that give the appearance of potentially influencing, what you wrote in the submitted work?

☐ Yes, the following relationships/conditions/circumstances are present (explain below):

☑ No other relationships/conditions/circumstances that present a potential conflict of interest

At the time of manuscript acceptance, journals will ask authors to confirm and, if necessary, update their disclosure statements. On occasion, journals may ask authors to disclose further information about reported relationships.

#### Section 6. Disclosure Statement

Based on the above disclosures, this form will automatically generate a disclosure statement, which will appear in the box below.

Jessica Reyes reports grants (to RFI) from the Bill & Melinda Gates Foundation and the Jacobs Institute for Molecular Engineering for Medicine (Caltech) , during the conduct of the study; .

#### Evaluation and Feedback

Please visit http://www.icmje.org/cgi-bin/feedback to provide feedback on your experience with completing this form.

### ICMJE Form for Disclosure of Potential Conflicts of Interest

#### Instructions

**The purpose of this form is to provide readers of your manuscript with information about your other interests that could influence how they receive and understand your work. The form is designed to be completed electronically and stored electronically. It contains programming that allows appropriate data display. Each author should submit a separate form and is responsible for the accuracy and completeness of the submitted information. The form is in six parts.**

#### 1. Identifying information.

#### 2. The work under consideration for publication.

This section asks for information about the work that you have submitted for publication. The time frame for this reporting is that of the work itself, from the initial conception and planning to the present. The requested information is about resources that you received, either directly or indirectly (via your institution), to enable you to complete the work. Checking "No" means that you did the work without receiving any financial support from any third party -- that is, the work was supported by funds from the same institution that pays your salary and that institution did not receive third-party funds with which to pay you. If you or your institution received funds from a third party to support the work, such as a government granting agency, charitable foundation or commercial sponsor, check "Yes".

#### 3. Relevant financial activities outside the submitted work.

This section asks about your financial relationships with entities in the bio-medical arena that could be perceived to influence, or that give the appearance of potentially influencing, what you wrote in the submitted work. You should disclose interactions with ANY entity that could be considered broadly relevant to the work. For example, if your article is about testing an epidermal growth factor receptor (EGFR) antagonist in lung cancer, you should report all associations with entities pursuing diagnostic or therapeutic strategies in cancer in general, not just in the area of EGFR or lung cancer.

Report all sources of revenue paid (or promised to be paid) directly to you or your institution on your behalf over the 36 months prior to submission of the work. This should include all monies from sources with relevance to the submitted work, not just monies from the entity that sponsored the research. Please note that your interactions with the work's sponsor that are outside the submitted work should also be listed here. If there is any question, it is usually better to disclose a relationship than not to do so.

For grants you have received for work outside the submitted work, you should disclose support ONLY from entities that could be perceived to be affected financially by the published work, such as drug companies, or foundations supported by entities that could be perceived to have a financial stake in the outcome. Public funding sources, such as government agencies, charitable foundations or academic institutions, need not be disclosed. For example, if a government agency sponsored a study in which you have been involved and drugs were provided by a pharmaceutical company, you need only list the pharmaceutical company.

#### 4. Intellectual Property.

This section asks about patents and copyrights, whether pending, issued, licensed and/or receiving royalties.

#### 5. Relationships not covered above.

Use this section to report other relationships or activities that readers could perceive to have influenced, or that give the appearance of potentially influencing, what you wrote in the submitted work.

##### Definitions.

**Entity:** government agency, foundation, commercial sponsor, academic institution, etc.
**Grant:** A grant from an entity, generally [but not always] paid to your organization
**Personal Fees:** Monies paid to you for services rendered, generally honoraria, royalties, or fees for consulting , lectures, speakers bureaus, expert testimony, employment, or other affiliations
**Non-Financial Support:** Examples include drugs/equipment supplied by the entity, travel paid by the entity, writing assistance, administrative support, etc**.**

**Other:** Anything not covered under the previous three boxes
**Pending:** The patent has been filed but not issued
**Issued:** The patent has been issued by the agency
**Licensed:** The patent has been licensed to an entity, whether earning royalties or not
**Royalties:** Funds are coming in to you or your institution due to your patent

### ICMJE Form for Disclosure of Potential Conflicts of Interest

#### Section 1. Identifying Information

1. Given Name (First Name)
Anna

2. Surname (Last Name)
Romano

3. Date
20-November-2020

4. Are you the corresponding author? ☐ Yes ☑ No

Corresponding Author's Name
Rustem F. Ismagilov

5. Manuscript Title
SARS-CoV-2 Viral Load Rises Gradually and to Moderate Levels in Some Humans

6. Manuscript Identifying Number (if you know it)
n/a

#### Section 2. The Work Under Consideration for Publication

Did you or your institution **at any time** receive payment or services from a third party (government, commercial, private foundation, etc.) for any aspect of the submitted work (including but not limited to grants, data monitoring board, study design, manuscript preparation, statistical analysis, etc.)?

Are there any relevant conflicts of interest? ☑ Yes ☐ No

If yes, please fill out the appropriate information below. If you have more than one entity press the "ADD" button to add a row. Excess rows can be removed by pressing the "X" button.

| Name of Institution/Company | Grant? | Personal Fees? | Non-Financial Support? | Other? | Comments |
|---|---|---|---|---|---|
| Bill & Melinda Gates Foundation | ☑ | ☐ | ☐ | ☐ | Grant to RFI |
| Jacobs Institute for Molecular Engineering for Medicine (Caltech) | ☑ | ☐ | ☐ | ☐ | Grant to RFI |

#### Section 3. Relevant financial activities outside the submitted work.

Place a check in the appropriate boxes in the table to indicate whether you have financial relationships (regardless of amount of compensation) with entities as described in the instructions. Use one line for each entity; add as many lines as you need by clicking the "Add +" box. You should report relationships that were **present during the 36 months prior to publication**.

Are there any relevant conflicts of interest? ☐ Yes ☑ No

#### Section 4. Intellectual Property -- Patents & Copyrights

Do you have any patents, whether planned, pending or issued, broadly relevant to the work? ☐ Yes ☑ No

ICMJE INTERNATIONAL COMMITTEE of MEDICAL JOURNAL EDITORS

### ICMJE Form for Disclosure of Potential Conflicts of Interest

#### Section 5. Relationships not covered above

Are there other relationships or activities that readers could perceive to have influenced, or that give the appearance of potentially influencing, what you wrote in the submitted work?

☐ Yes, the following relationships/conditions/circumstances are present (explain below):

☑ No other relationships/conditions/circumstances that present a potential conflict of interest

At the time of manuscript acceptance, journals will ask authors to confirm and, if necessary, update their disclosure statements. On occasion, journals may ask authors to disclose further information about reported relationships.

#### Section 6. Disclosure Statement

Based on the above disclosures, this form will automatically generate a disclosure statement, which will appear in the box below.

Mrs. Romano reports grants (to RFI) from Bill & Melinda Gates Foundation and the Jacobs Institute for Molecular Engineering for Medicine (Caltech), during the conduct of the study; .

#### Evaluation and Feedback

Please visit http://www.icmje.org/cgi-bin/feedback to provide feedback on your experience with completing this form.

INTERNATIONAL COMMITTEE of MEDICAL JOURNAL EDITORS

### ICMJE Form for Disclosure of Potential Conflicts of Interest

#### Instructions

**The purpose of this form is to provide readers of your manuscript with information about your other interests that could influence how they receive and understand your work. The form is designed to be completed electronically and stored electronically. It contains programming that allows appropriate data display. Each author should submit a separate form and is responsible for the accuracy and completeness of the submitted information. The form is in six parts.**

#### 1. Identifying information.

#### 2. The work under consideration for publication.

This section asks for information about the work that you have submitted for publication. The time frame for this reporting is that of the work itself, from the initial conception and planning to the present. The requested information is about resources that you received, either directly or indirectly (via your institution), to enable you to complete the work. Checking "No" means that you did the work without receiving any financial support from any third party -- that is, the work was supported by funds from the same institution that pays your salary and that institution did not receive third-party funds with which to pay you. If you or your institution received funds from a third party to support the work, such as a government granting agency, charitable foundation or commercial sponsor, check "Yes".

#### 3. Relevant financial activities outside the submitted work.

This section asks about your financial relationships with entities in the bio-medical arena that could be perceived to influence, or that give the appearance of potentially influencing, what you wrote in the submitted work. You should disclose interactions with ANY entity that could be considered broadly relevant to the work. For example, if your article is about testing an epidermal growth factor receptor (EGFR) antagonist in lung cancer, you should report all associations with entities pursuing diagnostic or therapeutic strategies in cancer in general, not just in the area of EGFR or lung cancer.

Report all sources of revenue paid (or promised to be paid) directly to you or your institution on your behalf over the 36 months prior to submission of the work. This should include all monies from sources with relevance to the submitted work, not just monies from the entity that sponsored the research. Please note that your interactions with the work's sponsor that are outside the submitted work should also be listed here. If there is any question, it is usually better to disclose a relationship than not to do so.

For grants you have received for work outside the submitted work, you should disclose support ONLY from entities that could be perceived to be affected financially by the published work, such as drug companies, or foundations supported by entities that could be perceived to have a financial stake in the outcome. Public funding sources, such as government agencies, charitable foundations or academic institutions, need not be disclosed. For example, if a government agency sponsored a study in which you have been involved and drugs were provided by a pharmaceutical company, you need only list the pharmaceutical company.

#### 4. Intellectual Property.

This section asks about patents and copyrights, whether pending, issued, licensed and/or receiving royalties.

#### 5. Relationships not covered above.

Use this section to report other relationships or activities that readers could perceive to have influenced, or that give the appearance of potentially influencing, what you wrote in the submitted work.

##### Definitions.

**Entity:** government agency, foundation, commercial sponsor, academic institution, etc.
**Grant:** A grant from an entity, generally [but not always] paid to your organization
**Personal Fees:** Monies paid to you for services rendered, generally honoraria, royalties, or fees for consulting , lectures, speakers bureaus, expert testimony, employment, or other affiliations
**Non-Financial Support:** Examples include drugs/equipment supplied by the entity, travel paid by the entity, writing assistance, administrative support, etc**.**

**Other:** Anything not covered under the previous three boxes
**Pending:** The patent has been filed but not issued
**Issued:** The patent has been issued by the agency
**Licensed:** The patent has been licensed to an entity, whether earning royalties or not
**Royalties:** Funds are coming in to you or your institution due to your patent

### ICMJE Form for Disclosure of Potential Conflicts of Interest

#### Section 1. Identifying Information

1. Given Name (First Name)
Emily

2. Surname (Last Name)
Savela

3. Date
20-November-2020

4. Are you the corresponding author? ☐ Yes ☑ No

Corresponding Author's Name
Rustem F. Ismagilov

5. Manuscript Title
SARS-CoV-2 Viral Load Rises Gradually and to Moderate Levels in Some Humans

6. Manuscript Identifying Number (if you know it)
n/a

#### Section 2. The Work Under Consideration for Publication

Did you or your institution **at any time** receive payment or services from a third party (government, commercial, private foundation, etc.) for any aspect of the submitted work (including but not limited to grants, data monitoring board, study design, manuscript preparation, statistical analysis, etc.)?

Are there any relevant conflicts of interest? ☑ Yes ☐ No

If yes, please fill out the appropriate information below. If you have more than one entity press the "ADD" button to add a row. Excess rows can be removed by pressing the "X" button.

| Name of Institution/Company | Grant? | Personal Fees? | Non-Financial Support? | Other? | Comments |
|---|---|---|---|---|---|
| Bill and Melinda Gates Foundation | ☑ | ☐ | ☐ | ☐ | Grant to RFI |
| Jacobs Institute for Molecular Engineering for Medicine (Caltech) | ☑ | ☐ | ☐ | ☐ | Grant to RFI |

#### Section 3. Relevant financial activities outside the submitted work.

Place a check in the appropriate boxes in the table to indicate whether you have financial relationships (regardless of amount of compensation) with entities as described in the instructions. Use one line for each entity; add as many lines as you need by clicking the "Add +" box. You should report relationships that were **present during the 36 months prior to publication**.

Are there any relevant conflicts of interest? ☐ Yes ☑ No

#### Section 4. Intellectual Property -- Patents & Copyrights

Do you have any patents, whether planned, pending or issued, broadly relevant to the work? ☐ Yes ☑ No

ICMJE INTERNATIONAL COMMITTEE of MEDICAL JOURNAL EDITORS

### ICMJE Form for Disclosure of Potential Conflicts of Interest

#### Section 5. Relationships not covered above

Are there other relationships or activities that readers could perceive to have influenced, or that give the appearance of potentially influencing, what you wrote in the submitted work?

☐ Yes, the following relationships/conditions/circumstances are present (explain below):

☑ No other relationships/conditions/circumstances that present a potential conflict of interest

At the time of manuscript acceptance, journals will ask authors to confirm and, if necessary, update their disclosure statements. On occasion, journals may ask authors to disclose further information about reported relationships.

#### Section 6. Disclosure Statement

Based on the above disclosures, this form will automatically generate a disclosure statement, which will appear in the box below.

Ms. Savela reports grants (to RFI) from the Bill & Melinda Gates Foundation and the Jacobs Institute for Molecular Engineering for Medicine (Caltech) , during the conduct of the study; .

#### Evaluation and Feedback

Please visit http://www.icmje.org/cgi-bin/feedback to provide feedback on your experience with completing this form.

ICMJE INTERNATIONAL COMMITTEE of MEDICAL JOURNAL EDITORS

### ICMJE Form for Disclosure of Potential Conflicts of Interest

#### Instructions

**The purpose of this form is to provide readers of your manuscript with information about your other interests that could influence how they receive and understand your work. The form is designed to be completed electronically and stored electronically. It contains programming that allows appropriate data display. Each author should submit a separate form and is responsible for the accuracy and completeness of the submitted information. The form is in six parts.**

#### 1. Identifying information.

#### 2. The work under consideration for publication.

This section asks for information about the work that you have submitted for publication. The time frame for this reporting is that of the work itself, from the initial conception and planning to the present. The requested information is about resources that you received, either directly or indirectly (via your institution), to enable you to complete the work. Checking "No" means that you did the work without receiving any financial support from any third party -- that is, the work was supported by funds from the same institution that pays your salary and that institution did not receive third-party funds with which to pay you. If you or your institution received funds from a third party to support the work, such as a government granting agency, charitable foundation or commercial sponsor, check "Yes".

#### 3. Relevant financial activities outside the submitted work.

This section asks about your financial relationships with entities in the bio-medical arena that could be perceived to influence, or that give the appearance of potentially influencing, what you wrote in the submitted work. You should disclose interactions with ANY entity that could be considered broadly relevant to the work. For example, if your article is about testing an epidermal growth factor receptor (EGFR) antagonist in lung cancer, you should report all associations with entities pursuing diagnostic or therapeutic strategies in cancer in general, not just in the area of EGFR or lung cancer.

Report all sources of revenue paid (or promised to be paid) directly to you or your institution on your behalf over the 36 months prior to submission of the work. This should include all monies from sources with relevance to the submitted work, not just monies from the entity that sponsored the research. Please note that your interactions with the work's sponsor that are outside the submitted work should also be listed here. If there is any question, it is usually better to disclose a relationship than not to do so.

For grants you have received for work outside the submitted work, you should disclose support ONLY from entities that could be perceived to be affected financially by the published work, such as drug companies, or foundations supported by entities that could be perceived to have a financial stake in the outcome. Public funding sources, such as government agencies, charitable foundations or academic institutions, need not be disclosed. For example, if a government agency sponsored a study in which you have been involved and drugs were provided by a pharmaceutical company, you need only list the pharmaceutical company.

#### 4. Intellectual Property.

This section asks about patents and copyrights, whether pending, issued, licensed and/or receiving royalties.

#### 5. Relationships not covered above.

Use this section to report other relationships or activities that readers could perceive to have influenced, or that give the appearance of potentially influencing, what you wrote in the submitted work.

##### Definitions.

**Entity:** government agency, foundation, commercial sponsor, academic institution, etc.
**Grant:** A grant from an entity, generally [but not always] paid to your organization
**Personal Fees:** Monies paid to you for services rendered, generally honoraria, royalties, or fees for consulting , lectures, speakers bureaus, expert testimony, employment, or other affiliations
**Non-Financial Support:** Examples include drugs/equipment supplied by the entity, travel paid by the entity, writing assistance, administrative support, etc**.**

**Other:** Anything not covered under the previous three boxes
**Pending:** The patent has been filed but not issued
**Issued:** The patent has been issued by the agency
**Licensed:** The patent has been licensed to an entity, whether earning royalties or not
**Royalties:** Funds are coming in to you or your institution due to your patent

### ICMJE Form for Disclosure of Potential Conflicts of Interest

#### Section 1. Identifying Information

1. Given Name (First Name)
Natasha

2. Surname (Last Name)
Shelby

3. Date
16-November-2020

4. Are you the corresponding author? ☐ Yes ☑ No

Corresponding Author's Name
Rustem F. Ismagilov

5. Manuscript Title
SARS-CoV-2 Viral Load Rises Gradually and to Moderate Levels in Some Humans

6. Manuscript Identifying Number (if you know it)
n/a

#### Section 2. The Work Under Consideration for Publication

Did you or your institution **at any time** receive payment or services from a third party (government, commercial, private foundation, etc.) for any aspect of the submitted work (including but not limited to grants, data monitoring board, study design, manuscript preparation, statistical analysis, etc.)?

Are there any relevant conflicts of interest? ☑ Yes ☐ No

If yes, please fill out the appropriate information below. If you have more than one entity press the "ADD" button to add a row. Excess rows can be removed by pressing the "X" button.

| Name of Institution/Company | Grant? | Personal Fees? | Non-Financial Support? | Other? | Comments |
|---|---|---|---|---|---|
| Bill & Melinda Gates Foundation | ☑ | ☐ | ☐ | ☐ | Grant to RFI |
| Jacobs Institute for Molecular Engineering for Medicine (Caltech) | ☑ | ☐ | ☐ | ☐ | Grant to RFI |

#### Section 3. Relevant financial activities outside the submitted work.

Place a check in the appropriate boxes in the table to indicate whether you have financial relationships (regardless of amount of compensation) with entities as described in the instructions. Use one line for each entity; add as many lines as you need by clicking the "Add +" box. You should report relationships that were **present during the 36 months prior to publication**.

Are there any relevant conflicts of interest? ☐ Yes ☑ No

#### Section 4. Intellectual Property -- Patents & Copyrights

Do you have any patents, whether planned, pending or issued, broadly relevant to the work? ☐ Yes ☑ No

INTERNATIONAL COMMITTEE of MEDICAL JOURNAL EDITORS

### ICMJE Form for Disclosure of Potential Conflicts of Interest

#### Section 5. Relationships not covered above

Are there other relationships or activities that readers could perceive to have influenced, or that give the appearance of potentially influencing, what you wrote in the submitted work?

☐ Yes, the following relationships/conditions/circumstances are present (explain below):

☑ No other relationships/conditions/circumstances that present a potential conflict of interest

At the time of manuscript acceptance, journals will ask authors to confirm and, if necessary, update their disclosure statements. On occasion, journals may ask authors to disclose further information about reported relationships.

#### Section 6. Disclosure Statement

Based on the above disclosures, this form will automatically generate a disclosure statement, which will appear in the box below.

Dr. Shelby reports grants (to RFI) from Bill & Melinda Gates Foundation and the Jacobs Institute for Molecular Engineering for Medicine (Caltech), during the conduct of the study; .

#### Evaluation and Feedback

Please visit http://www.icmje.org/cgi-bin/feedback to provide feedback on your experience with completing this form.

### ICMJE Form for Disclosure of Potential Conflicts of Interest

#### Instructions

**The purpose of this form is to provide readers of your manuscript with information about your other interests that could influence how they receive and understand your work. The form is designed to be completed electronically and stored electronically. It contains programming that allows appropriate data display. Each author should submit a separate form and is responsible for the accuracy and completeness of the submitted information. The form is in six parts.**

##### 1. Identifying information.

##### 2. The work under consideration for publication.

This section asks for information about the work that you have submitted for publication. The time frame for this reporting is that of the work itself, from the initial conception and planning to the present. The requested information is about resources that you received, either directly or indirectly (via your institution), to enable you to complete the work. Checking "No" means that you did the work without receiving any financial support from any third party -- that is, the work was supported by funds from the same institution that pays your salary and that institution did not receive third-party funds with which to pay you. If you or your institution received funds from a third party to support the work, such as a government granting agency, charitable foundation or commercial sponsor, check "Yes".

##### 3. Relevant financial activities outside the submitted work.

This section asks about your financial relationships with entities in the bio-medical arena that could be perceived to influence, or that give the appearance of potentially influencing, what you wrote in the submitted work. You should disclose interactions with ANY entity that could be considered broadly relevant to the work. For example, if your article is about testing an epidermal growth factor receptor (EGFR) antagonist in lung cancer, you should report all associations with entities pursuing diagnostic or therapeutic strategies in cancer in general, not just in the area of EGFR or lung cancer.

Report all sources of revenue paid (or promised to be paid) directly to you or your institution on your behalf over the 36 months prior to submission of the work. This should include all monies from sources with relevance to the submitted work, not just monies from the entity that sponsored the research. Please note that your interactions with the work's sponsor that are outside the submitted work should also be listed here. If there is any question, it is usually better to disclose a relationship than not to do so.

For grants you have received for work outside the submitted work, you should disclose support ONLY from entities that could be perceived to be affected financially by the published work, such as drug companies, or foundations supported by entities that could be perceived to have a financial stake in the outcome. Public funding sources, such as government agencies, charitable foundations or academic institutions, need not be disclosed. For example, if a government agency sponsored a study in which you have been involved and drugs were provided by a pharmaceutical company, you need only list the pharmaceutical company.

##### 4. Intellectual Property.

This section asks about patents and copyrights, whether pending, issued, licensed and/or receiving royalties.

##### 5. Relationships not covered above.

Use this section to report other relationships or activities that readers could perceive to have influenced, or that give the appearance of potentially influencing, what you wrote in the submitted work.

##### Definitions.

**Entity:** government agency, foundation, commercial sponsor, academic institution, etc.
**Grant:** A grant from an entity, generally [but not always] paid to your organization
**Personal Fees:** Monies paid to you for services rendered, generally honoraria, royalties, or fees for consulting , lectures, speakers bureaus, expert testimony, employment, or other affiliations
**Non-Financial Support:** Examples include drugs/equipment supplied by the entity, travel paid by the entity, writing assistance, administrative support, etc**.**

**Other:** Anything not covered under the previous three boxes
**Pending:** The patent has been filed but not issued
**Issued:** The patent has been issued by the agency
**Licensed:** The patent has been licensed to an entity, whether earning royalties or not
**Royalties:** Funds are coming in to you or your institution due to your patent

INTERNATIONAL COMMITTEE of MEDICAL JOURNAL EDITORS

### ICMJE Form for Disclosure of Potential Conflicts of Interest

#### Section 1. Identifying Information

1. Given Name (First Name)
Colten

2. Surname (Last Name)
Tognazzini

3. Date
22-November-2020

4. Are you the corresponding author? ☐ Yes ☑ No

Corresponding Author's Name
Rustem F. Ismagilov

5. Manuscript Title
SARS-CoV-2 Viral Load Rises Gradually and to Moderate Levels in Some Humans

6. Manuscript Identifying Number (if you know it)

#### Section 2. The Work Under Consideration for Publication

Did you or your institution **at any time** receive payment or services from a third party (government, commercial, private foundation, etc.) for any aspect of the submitted work (including but not limited to grants, data monitoring board, study design, manuscript preparation, statistical analysis, etc.)?

Are there any relevant conflicts of interest? ☐ Yes ☑ No

#### Section 3. Relevant financial activities outside the submitted work.

Place a check in the appropriate boxes in the table to indicate whether you have financial relationships (regardless of amount of compensation) with entities as described in the instructions. Use one line for each entity; add as many lines as you need by clicking the "Add +" box. You should report relationships that were **present during the 36 months prior to publication**.

Are there any relevant conflicts of interest? ☐ Yes ☑ No

#### Section 4. Intellectual Property -- Patents & Copyrights

Do you have any patents, whether planned, pending or issued, broadly relevant to the work? ☐ Yes ☑ No

### ICMJE Form for Disclosure of Potential Conflicts of Interest

#### Section 5. Relationships not covered above

Are there other relationships or activities that readers could perceive to have influenced, or that give the appearance of potentially influencing, what you wrote in the submitted work?

☐ Yes, the following relationships/conditions/circumstances are present (explain below):

☑ No other relationships/conditions/circumstances that present a potential conflict of interest

At the time of manuscript acceptance, journals will ask authors to confirm and, if necessary, update their disclosure statements. On occasion, journals may ask authors to disclose further information about reported relationships.

#### Section 6. Disclosure Statement

Based on the above disclosures, this form will automatically generate a disclosure statement, which will appear in the box below.

Dr. Tognazzini has nothing to disclose.

#### Evaluation and Feedback

Please visit http://www.icmje.org/cgi-bin/feedback to provide feedback on your experience with completing this form.

### ICMJE Form for Disclosure of Potential Conflicts of Interest

#### Instructions

**The purpose of this form is to provide readers of your manuscript with information about your other interests that could influence how they receive and understand your work. The form is designed to be completed electronically and stored electronically. It contains programming that allows appropriate data display. Each author should submit a separate form and is responsible for the accuracy and completeness of the submitted information. The form is in six parts.**

#### 1. Identifying information.

#### 2. The work under consideration for publication.

This section asks for information about the work that you have submitted for publication. The time frame for this reporting is that of the work itself, from the initial conception and planning to the present. The requested information is about resources that you received, either directly or indirectly (via your institution), to enable you to complete the work. Checking "No" means that you did the work without receiving any financial support from any third party -- that is, the work was supported by funds from the same institution that pays your salary and that institution did not receive third-party funds with which to pay you. If you or your institution received funds from a third party to support the work, such as a government granting agency, charitable foundation or commercial sponsor, check "Yes".

#### 3. Relevant financial activities outside the submitted work.

This section asks about your financial relationships with entities in the bio-medical arena that could be perceived to influence, or that give the appearance of potentially influencing, what you wrote in the submitted work. You should disclose interactions with ANY entity that could be considered broadly relevant to the work. For example, if your article is about testing an epidermal growth factor receptor (EGFR) antagonist in lung cancer, you should report all associations with entities pursuing diagnostic or therapeutic strategies in cancer in general, not just in the area of EGFR or lung cancer.

Report all sources of revenue paid (or promised to be paid) directly to you or your institution on your behalf over the 36 months prior to submission of the work. This should include all monies from sources with relevance to the submitted work, not just monies from the entity that sponsored the research. Please note that your interactions with the work's sponsor that are outside the submitted work should also be listed here. If there is any question, it is usually better to disclose a relationship than not to do so.

For grants you have received for work outside the submitted work, you should disclose support ONLY from entities that could be perceived to be affected financially by the published work, such as drug companies, or foundations supported by entities that could be perceived to have a financial stake in the outcome. Public funding sources, such as government agencies, charitable foundations or academic institutions, need not be disclosed. For example, if a government agency sponsored a study in which you have been involved and drugs were provided by a pharmaceutical company, you need only list the pharmaceutical company.

#### 4. Intellectual Property.

This section asks about patents and copyrights, whether pending, issued, licensed and/or receiving royalties.

#### 5. Relationships not covered above.

Use this section to report other relationships or activities that readers could perceive to have influenced, or that give the appearance of potentially influencing, what you wrote in the submitted work.

##### Definitions.

**Entity:** government agency, foundation, commercial sponsor, academic institution, etc.
**Grant:** A grant from an entity, generally [but not always] paid to your organization
**Personal Fees:** Monies paid to you for services rendered, generally honoraria, royalties, or fees for consulting , lectures, speakers bureaus, expert testimony, employment, or other affiliations
**Non-Financial Support:** Examples include drugs/equipment supplied by the entity, travel paid by the entity, writing assistance, administrative support, etc**.**

**Other:** Anything not covered under the previous three boxes
**Pending:** The patent has been filed but not issued
**Issued:** The patent has been issued by the agency
**Licensed:** The patent has been licensed to an entity, whether earning royalties or not
**Royalties:** Funds are coming in to you or your institution due to your patent

INTERNATIONAL COMMITTEE of MEDICAL JOURNAL EDITORS

### ICMJE Form for Disclosure of Potential Conflicts of Interest

#### Section 1. Identifying Information

1. Given Name (First Name)
Alexander

2. Surname (Last Name)
Winnett

3. Date
20-November-2020

4. Are you the corresponding author? ☐ Yes ☑ No

Corresponding Author's Name
Rustem F. Ismagilov

5. Manuscript Title
SARS-CoV-2 Viral Load Rises Gradually and to Moderate Levels in Some Humans

6. Manuscript Identifying Number (if you know it)

#### Section 2. The Work Under Consideration for Publication

Did you or your institution **at any time** receive payment or services from a third party (government, commercial, private foundation, etc.) for any aspect of the submitted work (including but not limited to grants, data monitoring board, study design, manuscript preparation, statistical analysis, etc.)?

Are there any relevant conflicts of interest? ☑ Yes ☐ No

If yes, please fill out the appropriate information below. If you have more than one entity press the "ADD" button to add a row. Excess rows can be removed by pressing the "X" button.

| Name of Institution/Company | Grant? | Personal Fees? | Non-Financial Support? | Other? | Comments |
|---|---|---|---|---|---|
| Bill & Melinda Gates Foundation | ☑ | ☐ | ☐ | ☐ | Grant to RFI |
| Jacobs Institute for Molecular Engineering for Medicine (Caltech) | ☑ | ☐ | ☐ | ☐ | Grant to RFI |
| National Institutes of Health NIGMS Predoctoral Training Grant (GM008042) | ☐ | ☐ | ☐ | ☑ | Fellowship to AW |
| UCLA Geffen Fellowship | ☐ | ☐ | ☐ | ☑ | Fellowship to AW |

#### Section 3. Relevant financial activities outside the submitted work.

Place a check in the appropriate boxes in the table to indicate whether you have financial relationships (regardless of amount of compensation) with entities as described in the instructions. Use one line for each entity; add as many lines as you need by clicking the "Add +" box. You should report relationships that were **present during the 36 months prior to publication**.

Are there any relevant conflicts of interest? ☐ Yes ☑ No

### ICMJE Form for Disclosure of Potential Conflicts of Interest

#### Section 4. Intellectual Property -- Patents & Copyrights

Do you have any patents, whether planned, pending or issued, broadly relevant to the work? ☐ Yes ☑ No

#### Section 5. Relationships not covered above

Are there other relationships or activities that readers could perceive to have influenced, or that give the appearance of potentially influencing, what you wrote in the submitted work?

☐ Yes, the following relationships/conditions/circumstances are present (explain below):

☑ No other relationships/conditions/circumstances that present a potential conflict of interest

At the time of manuscript acceptance, journals will ask authors to confirm and, if necessary, update their disclosure statements. On occasion, journals may ask authors to disclose further information about reported relationships.

#### Section 6. Disclosure Statement

Based on the above disclosures, this form will automatically generate a disclosure statement, which will appear in the box below.

Dr. Winnett reports grants (to RFI) from the Bill & Melinda Gates Foundation and the Jacobs Institute for Molecular Engineering for Medicine (Caltech), and a National Institutes of Health NIGMS Predoctoral Training Grant (GM008042) and a UCLA Geffen Fellowship, during the conduct of the study; .

#### Evaluation and Feedback

Please visit http://www.icmje.org/cgi-bin/feedback to provide feedback on your experience with completing this form.
