## Supplemental Information for "SARS-CoV-2 Viral Load in Saliva Rises Gradually and to Moderate Levels in Some Humans"

**Contents:**

Supplemental Tables 1-2
Supplemental Figures 1-2
Supplemental References
Contributions of Non-Corresponding Authors

**TABLE S1. Pertinent Demographic and Health Information for Study Participants.**

| | Household Members | | | |
|---|---|---|---|---|
| | **Parent-1** | **Parent-2** | **Sibling-1** | **Sibling-2** |
| **Age Range (Years)** | 50-60 | 50-60 | 18-25 | 18-25 |
| **Race, Ethnicity** | White, Non-Hispanic | White, Non-Hispanic | White, Non-Hispanic | White, Non-Hispanic |
| **Self-Described Health Status** | “Excellent” | “Excellent” | “Excellent” | “Excellent” |
| **Reported Medical Conditions** | None | None | None | None |

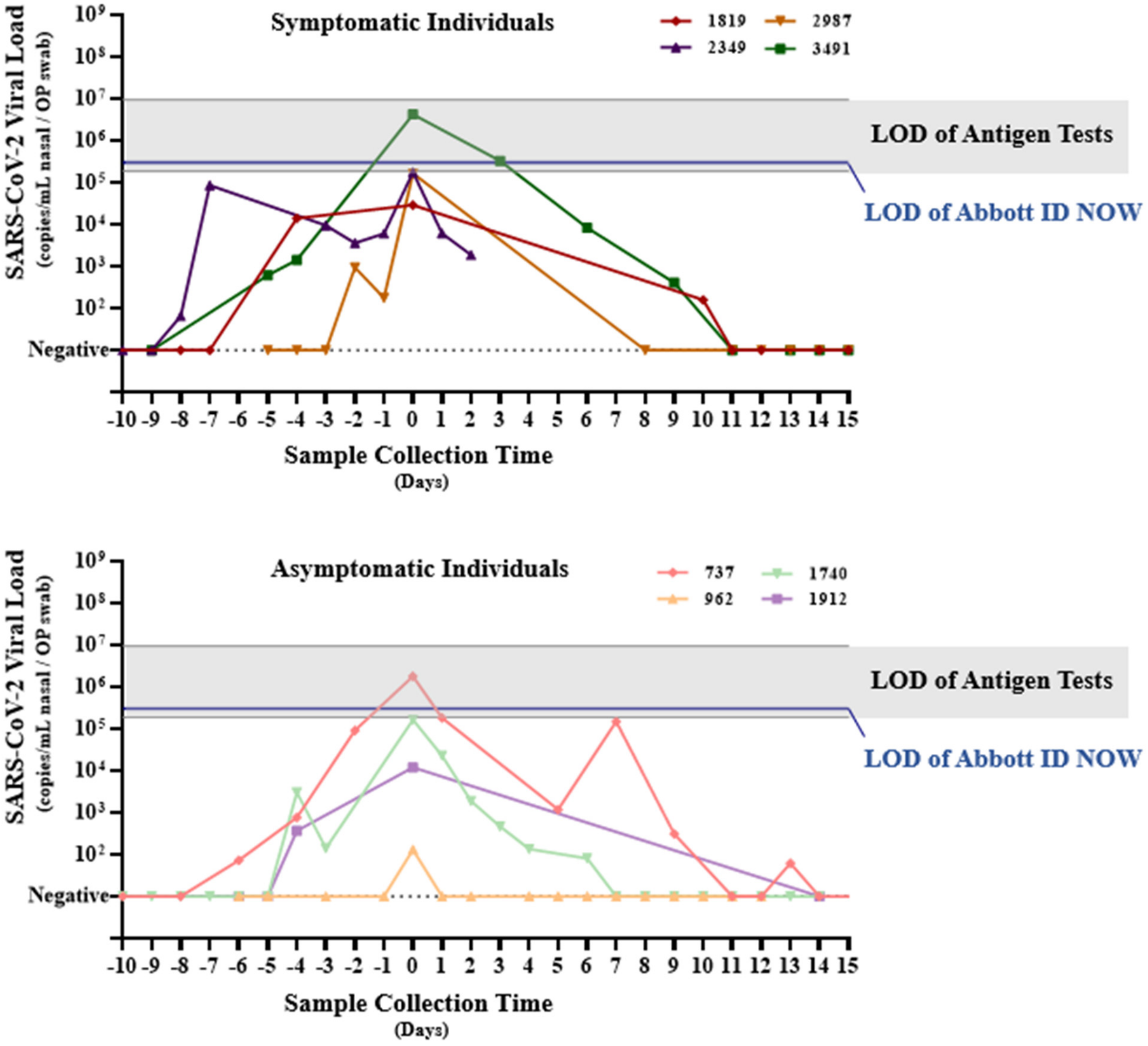


**FIGURE S1. Re-analysis of Select Data from Kissler et al. (2020).**[1] Cycle threshold values from the eight individuals who had at least 3 measurements from anterior nares and oropharyngeal (OP) swabs during the pre-peak period of infection were converted to viral load and replotted. Viral loads are plotted with peak viral load assigned to Day 0. A) Viral load profiles of individuals who became symptomatic and B) Viral load profiles of individuals who did not report symptoms. Horizontal blue line on each plot depicts the limit of detection (LOD) of Abbott ID NOW ($3x10^5$ copies/mL from U.S. FDA SARS-CoV-2 Reference Panel Comparative testing data).[2] Horizontal grey bar depicts the range of LODs for commercial antigen tests ($1.90x10^5$ copies/mL to $9.33x10^6$ copies/mL; see Table S2).

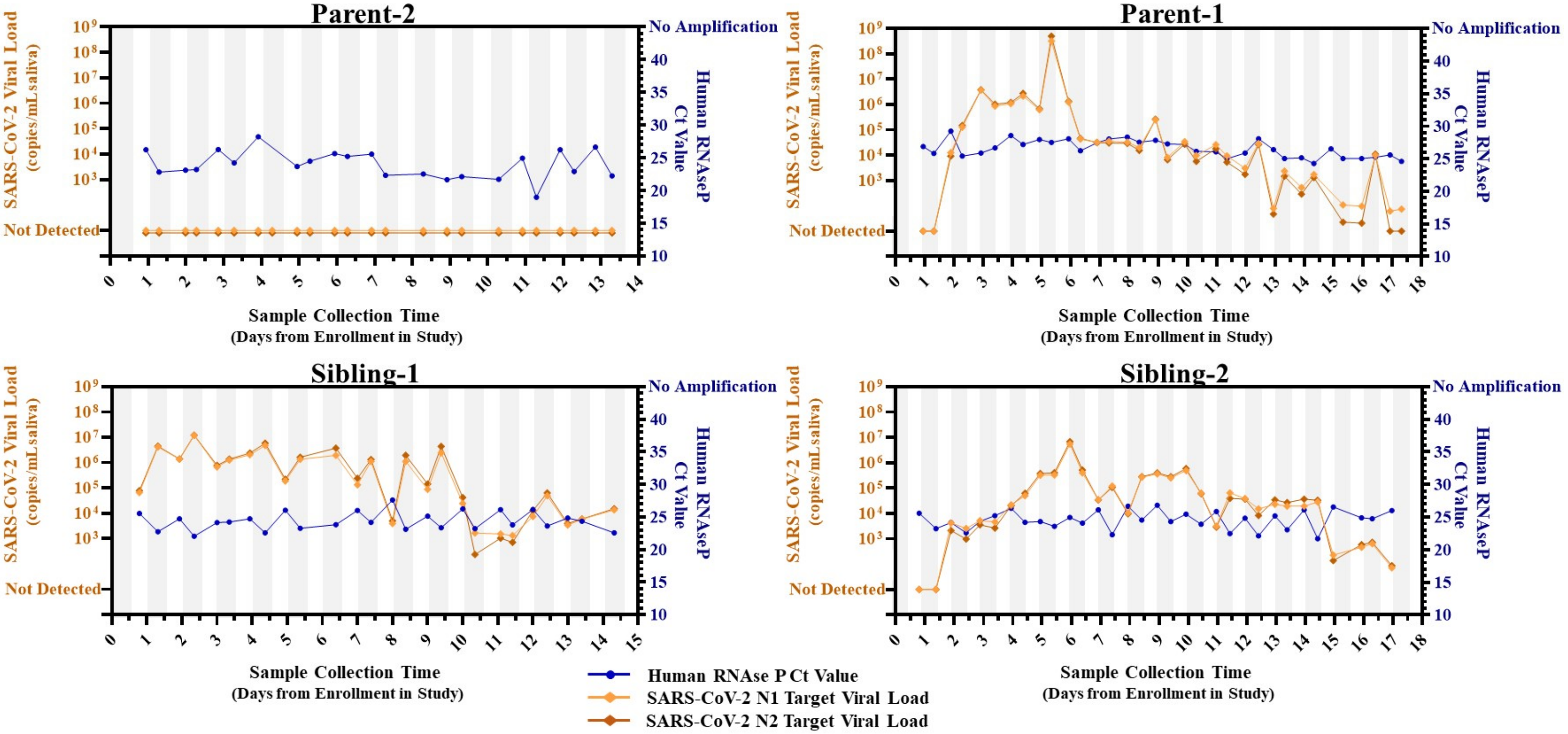


**FIGURE S2. Human RNase P Ct values and SARS-CoV-2 viral load measurements from RT-qPCR for all household contacts.**

**TABLE S2. Summary of Reported Limits of Detection (LOD) for Rapid Point of Care SARS-CoV-2 Antigen Tests.** Information on limit of detection was collected from instructions for use (IFU) and from published literature that reported LOD in units of copies/mL or could be unambiguously converted to copies/mL.

| Company | Product | Limit of Detection (copies/mL) | Reference | Explanation |
|---|---|---|---|---|
| **Quidel** | **Sofia SARS Antigen FIA** | 6.0x$10^{6}$ | Arnaout et al.[3] | Estimated from US FDA Emergency Use Authorization Instructions For Use |
| | | 1.36x$10^{6}$ | Pollack et al.[4] | In the US FDA Emergency Use Authorization Instructions For Use, the Limit of Detection is reported as Tissue Culture Infectious Dose 50 (TCID50). Pollack et al. determine a conversion from TCID50 per volume to mass of antigen, and mass of antigen per volume to RNA copies per volume. |
| **BD** | **Veritor™ Plus System for rapid COVID-19 (SARS-CoV-2) Test** | 8.07x$10^{5}$ | | |
| **Lumira Dx** | **COVID-19 SARS-CoV-2 Antigen Test** | 1.9x$10^{5}$ | Instructions for Use[5] | Estimated from US FDA Emergency Use Authorization Instructions For Use. The Limit of Detection is reported as TCID50 per volume. The lot of virus from BEI that was used for testing is reported in the IFU, and the certificate of analysis for that lot provides conversion from the TCID50 value to RNA copies per volume. |
| **Abbott** | **PANBIO™ COVID-19 Ag Rapid Test Device** | 7.94x$10^{5}$ | Albert et al.[6] | Albert et al report the results of samples tested by RT-qPCR (converted to viral load) and the stated POC antigen test to define the Limit of Detection. |
| | | 3.55x$10^{6}$ | Corman et al.[7] | Corman et al. report the results of samples tested by RT-qPCR (converted to viral load) and the stated POC antigen test to define the Limit of Detection. Two tests with particularly poor analytical limit of detection (RapidGEN Biocredit COVID-19 Ag Test, LOD = 1.58x$10^{10}$ and Coris Bioconcept COVID 19 Ag Respi-Strip, LOD=2.88x$10^{7}$) were excluded from this table as not representative. |
| **Healgen** | **Coronavirus Ag Rapid Test Cassette (Swab)** | 2.34x$10^{6}$ | | |
| **R-Biopharm** | **RIDA QUICK SARS-CoV-2 Ag** | 2.09x$10^{6}$ | | |
| **Nal-vonminden** | **NADAL COVID19-Ag** | 9.33x$10^{6}$ | | |
| **Roche** | **SD Biosensor SARS-CoV-2 Rapid Antigen Test** | 6.03x$10^{6}$ | | |

**NON-CORRESPONDING AUTHOR CONTRIBUTIONS**

AW - Collaborated with MMC, NS, RFI, YG, MF on initial study design and recruitment strategies; co-wrote IRB protocol and informed consent with MMC and NS; co-wrote enrollment questionnaire with NS and JJ; co-wrote participant informational sheets with NS and JAR; reagents and supplies acquisition; funding acquisition; assisted in sample logging system implementation with JTB; developed laboratory sample processing workflow with AER and MMC; performed nucleic acid extraction, and RT-qPCR; co-wrote and edited the manuscript; prepared Figure 1, Figure S1 with RA, Figure S2, Table S1 with NS, JAR and JJ, and Table S2.

MMC – Collaborated with AW, NS, RFI, YG, MF on initial study design and recruitment strategies; co-wrote IRB protocol and informed consent with AW and NS; assisted in the writing of the enrollment questionnaire; developed laboratory sample processing workflow with AW and AER; contributed to the analyses of the RT-qPCR and RT-ddPCR data; validated the LOD of the assay; funding acquisition; co-wrote and edited the manuscript.

NS - Study administrator; collaborated with AW, MMC, RFI, YG, MF on initial study design and recruitment strategies; co-wrote IRB protocol and informed consent with AW and MMC; co-wrote enrollment questionnaire with AW and JJ; co-wrote participant informational sheets with AW and JAR; enrolled and maintained study participants with JAR; reagents and supplies acquisition; co-wrote and edited the manuscript.

AER -Developed laboratory sample processing workflow with AW and MMC; reagents and supplies acquisition; developed and validated method for RT-qPCR and RT-digital droplet PCR analysis of extracted saliva samples; performed RT-qPCR and RT-digital droplet PCR;

JAR –Study coordinator; collaborated with NS, AW, RFI on recruitment strategies, translated study materials into Spanish, co-wrote informational sheets with AW and NS, enrolled and maintained study participants with NS.

JJ – Contributed to study design and study organization and implementation with NS and JAR; co-wrote enrollment questionnaire with NS and AW. Provided quality control and curation of participant data.

MP – Prepared participant sample collection materials, helped with supplies acquisition, and performed sample processing (RT-qPCR).

ES – Major contributor to biosafety SOPs and setting up lab workflow. Conducted biosafety training. Organized and coordinated lab work for the team. Performed sample processing including specimen logging and RT-qPCR. Minor contributor to validation of LOD of the assay.

JTB – Created specimen tracking database to aid in specimen logging and tracking.

RA – Provided literature review and processed data used in Figure S1.

CT – Coordinated the recruitment efforts at PPHD with case investigators and contact tracers.

MF – Co-investigator; collaborated with AW, MMC, NS, YG, RFI on study design and recruitment strategies; provided guidance and expertise on SARS-CoV-2 epidemiology and local trends.

YG – Co-investigator; collaborated with AW, MMC, NS, MF, RFI on study design and recruitment strategies; provided guidance and expertise on SARS-CoV-2 epidemiology and local trends.
